# Ranking Pretrained Speech Embeddings in Parkinson’s Disease Detection: Does Wav2Vec 2.0 Outperform its 1.0 Version Across Speech Modes and Languages?

**DOI:** 10.1101/2025.01.29.25321319

**Authors:** Ondrej Klempir, Adela Skryjova, Ales Tichopad, Radim Krupicka

## Abstract

Speech and language technologies are effective tools for identifying the distinct speech changes associated with Parkinson’s disease (PD), enabling earlier and more accurate diagnosis. Recent advancements in self-supervised speech pretraining, particularly with Wav2Vec models, have demonstrated superior performance over traditional feature extraction methods. While Wav2Vec 2.0 has been successfully utilized for PD detection, a rigorous quantitative comparison with Wav2Vec 1.0 is needed to comprehensively evaluate its advantages, limitations, and applicability across different speech modes in PD. This study presents a systematic comparison of Wav2Vec 1.0 and Wav2Vec 2.0 embeddings across three multilingual datasets using various classification approaches in classifying normal (healthy controls; HC) and PD speech. Additionally, both Wav2Vec versions were benchmarked against traditional baseline features across diverse linguistic contexts, including spontaneous speech, non-spontaneous speech, and isolated vowels. A multicriteria TOPSIS approach was employed to rank feature extraction methods, revealing that the Wav2Vec 2.0 consistently excelled across all speech modes, with its first transformer layer demonstrating the best performance for contextual tasks (read text and monologue) and its feature extractor performing best in vowel-based classification. In contrast, the Wav2Vec 1.0, while generally outperformed by the Wav2Vec 2.0, still provided a faster alternative with competitive performance in contextual tasks, highlighting its potential for specific applications, such as federated learning. This comparative analysis furthermore underscores the strengths of each Wav2Vec architecture and informs their optimal use in PD detection.

## Introduction

The integration of artificial intelligence (AI) into healthcare offers an immense potential for enhancing disease diagnostics [1]. Within AI, speech and language technologies have emerged as valuable tools for investigating language production in patients with Parkinson’s disease (PD). PD is associated with a variety of speech-related impairments, including dysarthria and language disorders [2]. Early detection of subtle dysarthria patterns in the prodromal stage of PD could enable earlier intervention with disease-modifying treatments [3].

Speech recordings play a critical role in identifying speech biomarkers and are evaluated using diverse acoustic analysis methods [2]. Detection of PD based on audio signals relies on contrasting signals obtained from healthy controls (HC) compared to signals from PD patients. Current advanced methods emphasize high-performing self-supervised deep learning. Recently, Nasersharif and Namvarpour categorized self-supervised models for robust speech representation learning into four groups [4]: recurrent models (typically employing modules like Long Short-Term Memory), Convolutional Neural Network (CNN)-based models, transformer-based models, and hybrid models that combine CNNs with transformers.

This study focuses on pre-trained model embeddings, particularly on the Wav2Vec (referred to as wav2vec1, wav2vec 1.0, or w2v1) [5], a CNN-based model that employs multi-layer CNNs to encode speech utterances, and the Wav2Vec 2.0 (referred to as wav2vec2, wav2vec 2.0, or w2v2) [6], a representative of hybrid models according to Nasersharif and Namvarpour’s [4]. For dysarthria detection, pre-trained embeddings have been shown to outperform traditional acoustic features [7]. Furthermore, Cai et al. [8], demonstrated the effectiveness of combining wav2vec 2.0 embeddings with machine learning models to classify normal and pathological voices, highlighting the benefits of audio augmentation. The Wav2Vec 2.0 has also shown promise in detecting prodromal speech patterns in PD [3]. Alongside the wav2vec 2.0, other digital architectures, such as HuBERT, TRILLsson, and Whisper, have also demonstrated superior performance in related classification tasks [3, 9, 10].

In addition to achieving a strong performance across various multilingual corpora and scenarios, the wav2vec 2.0 has the ability to filter out pathology-unrelated fluctuations in spontaneous speech [11]. This capability could facilitate broader practical implementation, as spontaneous speech is easier and more convenient to record in real-world settings. In contrast, classical approaches can struggle to effectively capture pathology-specific cues in such naturalistic speech contexts [11]. Another example of a recent comprehensive pipeline for analyzing speech and its linguistic properties in PD involved recording speech, transcribing it into text using Automatic Speech Recognition (ASR), and conducting further analyses such as part-of-speech tagging and syntactic complexity assessment [12]. ASR systems, including wav2vec-based ASR, are often utilized to assess speech intelligibility in PD [13, 14].

In our previous studies on the use of the wav2vec 1.0, we found that this model demonstrated a potential for developing a universal speech-based PD evaluation model as it outperformed traditional feature extraction methods in cross-database classification [15, 16]. Besides our own research, other studies investigating the wav2vec for the detection of PD have focused exclusively on the transformer-based wav2vec 2.0. To our best knowledge, the wav2vec is usually implicitly associated with wav2vec 2.0. However, it is critical to emphasize the existence of multiple versions, necessitating a thorough comparison to evaluate their respective advantages and limitations, particularly regarding their applicability across different speech modes in PD. This clarification aims to address and preclude potential misconceptions in the field.

We feel that the rapidly evolving scientific landscape would benefit from more focused and detailed comparison of the two versions of wav2vec for their uses in classifying normal versus PD speech. This study provides a direct comparison of the two embedding architectures w2v1 and w2v2 using three different datasets, and employs a wide range of classification methods. Further, the w2v are compared with traditional baseline features such as energy entropy or zero crossing rate. Beyond this, our aim was to explore the suitability of these models in various linguistic contexts, including spontaneous speech, non-spontaneous speech, and isolated vowels.

## Methods

### Speech corpuses

To demonstrate the proposed methods, address language differences, and evaluate the impact of various speech modes, we utilized audio recordings from three databases—an English database, a Spanish database, and a database focused on vowel recordings. In all the proposed modeling tasks, the target variable was consistently the prediction of PD based on the audio recordings (PD vs. HC).

#### English recordings

The dataset used for English recordings was the Mobile Device Voice Recordings at King’s College London (MDVR-KCL) [17]. This database includes high-quality audio recordings with a sampling rate of 44.1 kHz, featuring both read text and spontaneous dialogue. For the read text, participants were asked to recite a fixed passage, “The North Wind and the Sun”. Spontaneous dialogue was initiated by test administrators asking open-ended questions about topics such as local attractions, traffic, or personal interests. The dataset is slightly imbalanced, comprising of 21 HC and 16 participants with PD. In such cases, baseline accuracy—achieved by predicting all samples as HC—would yield 57%. Clinical scores for the participants include the Hoehn and Yahr (H&Y) scale, with a mean score of 2.6 ± 0.72, and the Unified Parkinson’s Disease Rating Scale (UPDRS) III, speech subscore (item 18), with a mean of 0.94 ± 0.93. Detailed demographic information for the participants is unavailable. This corpus features more complex speech tasks, and does not provide isolated vowels.

#### Spanish recordings

The Spanish dataset used in this study is PC-GITA, a well-known resource in PD research [18]. This dataset is balanced in terms of both sample size and gender distribution, consisting of 50 participants with PD (25 female, 25 male) and 50 HC (25 female, 25 male). For the PD group, the mean total UPDRS score was 37.7 ± 18.3, the mean UPDRS speech component was 1.3 ± 0.8, the mean H&Y scale was 2.2 ± 0.7, and the mean age was 61 ± 9.4 years. For the HC group, the mean age was 61 ± 9.5 years. The recordings had a sampling frequency of 44.1 kHz and included a variety of speech tasks, such as a read text, spontaneous monologues, and isolated vowels. For individuals with multiple recordings of the same task, only a single instance was selected for use in the proposed models, to prevent potential data leakage.

#### Sustained vowel /a/ recordings

For sustained vowel analysis, we utilized a U.S.-based dataset [19]. This dataset consists of personal, telephone-collected recordings of the sustained vowel /a/ in natural settings (8 kHz sampling frequency), including samples from 50 participants with specialist-diagnosed PD and 50 HC. After pre-processing, the final study population comprised of 40 individuals with PD and 41 HC. The recordings represent prolonged enunciations of the vowel /a/ captured using participants’ personal telephones. For the HC group, there were 16 males and 25 females with a mean age of 47.9 ± 14.5 years. For the PD group, there were 21 males and 19 females with a mean age of 66.6 ± 9.0 years and a mean H&Y score of 2.1 ± 0.4. Additionally, the vowel analysis was further extended using /a/ recordings from the PC-GITA dataset.

#### Traditional acoustic features

To establish a simple, straightforward, and easily interpretable baseline, we employed six traditional handcrafted acoustic features. These features were computed following the approach described in [20]. The feature extraction process involved two steps:

1. The audio signal was segmented into short-term, non-overlapping 50 ms frames. For each frame, six acoustic features were calculated (details can be found in Suppl. Methods 1): Energy Entropy (A), Energy (B), Zero Crossing Rate (C), Spectral Rolloff (D), Spectral Centroid (E), and Spectral Flux (F). This resulted in six feature sequences representing the entire audio signal.
2. For each of the six feature sequences, a descriptive statistic was computed (Table 1), producing a single summary value for each feature. These six summary values collectively represent the final feature set that characterizes the input speech signal.

**Table 1.** List of calculated traditional features.

| Feature | Statistic | Domain |
| --- | --- | --- |
| EnergyEntropy | std | time |
| SignalEnergy | std/mean | time |
| ZeroCrossingRate | std | time |
| SpectralRolloff | std | spectral |
| SpectralCentroid | std | spectral |
| SpectralFlux | std/mean | spectral |

### Speech pretrained embeddings

We focused on two architecture versions of the Wav2Vec, the 1.0 and 2.0 versions (Supplementary Fig. 1). Wav2Vec-based features are inherently non-interpretable but serve as universal representations, eliminating the need for handcrafted feature engineering. As a preprocessing step, all speech signals were resampled to 16 kHz using *torchaudio.transforms.Resample* [21]. Both Wav2Vec architectures were originally trained in a self-supervised manner, optimizing a contrastive loss function. We did not perform fine-tuning on the Wav2Vec architectures.

#### Wav2Vec 1.0

We utilized the original Wav2Vec model [5]. This is a CNN-based architecture that employs multi-layer convolutional neural networks to encode speech utterances. Wav2Vec 1.0 comprises of two cascaded CNN modules, a feature extractor and a feature aggregator. The feature extractor refines raw audio signals into preliminary representations, while the feature aggregator integrates these representations into higher-level latent variables that capture contextual semantic relationships among audio features [22].

For our analysis, we employed the “wav2vec_large” variant, which has an expanded capacity due to its larger context network, consisting of twelve layers [23]. This model was pre-trained on the LibriSpeech dataset, comprising 960 hours of English speech sampled at 16 kHz. We extracted representations from two points in the model: (1) the output of the feature extractor (FE) and (2) the combined output of the feature extractor and aggregator (FEA).

#### Wav2Vec 2.0

Wav2Vec 2.0 is a hybrid model that integrates CNNs with transformers. Each transformer layer processes the input through self-attention mechanisms and feed-forward networks, producing intermediate representations that encapsulate progressively more abstract and semantically rich information. As a result, the outputs from different transformer layers can encode diverse levels of information about the input audio, ranging from phonetic details to higher-level semantic patterns.

For our analysis, we employed the Wav2Vec 2.0 XLSR-53 model, a multilingual variant pre-trained on 53 languages, including English and Spanish [24, 25]. We focused on extracting representations from several key points in the CNN and transformer stack: the feature extractor, the first transformer layer and the last hidden layer. By analyzing these layers, we aimed to understand the evolution of speech representations across the network.

#### Embeddings aggregation

The obtained Wav2Vec representations were converted into fixed-sized, utterance-level feature vectors using mean aggregation. Vetráb and Gosztolya explored various aggregation techniques for Wav2Vec 2.0 deep embeddings, evaluating 11 different functions, and confirming that mean aggregation was an effective choice [26]. For Wav2Vec 1.0, the aggregation resulted in 512-dimensional vectors, while the Wav2Vec 2.0 dimensions varied depending on the selected layer, yielding either 1024-(transformer) or 512-dimensional (feature extractor) vectors. To further process the high-dimensional representations, we applied Principal Component Analysis (PCA) to decorrelate the feature space. From each Wav2Vec representation, we extracted the first 30 PCA components for subsequent analysis.

### Classification methods

The feature representations generated for each audio recording (w2v1, w2v2 and baselines) were used as input features for a variety of classification methods implemented in the Python library scikit-learn [27]. Specifically, we evaluated the following classifiers: Naive Bayes (NB; Gaussian NB), Support Vector Machine (SVM; with a linear kernel and probabilistic output), Decision Tree (DT; using default parameters), K-Nearest Neighbors (KNN; with n_neighbors=5), Logistic Regression (LR; with max_iter=1000), and Random Forest (RF; with n_estimators=100). Each classifier was initially trained using default or standard hyperparameter settings.

### Metrics and evaluation strategy

To assess the performance of the HC vs. PD classification models, we used the following metrics (True Positives, TP; True Negatives, TN; False Positives, FP; False Negatives, FN):

- *Accuracy:* Defined as 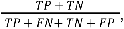 the accuracy metric measures the proportion of correctly classified instances in the dataset.

- *Sensitivity (Recall):* Calculated as 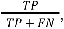 sensitivity evaluates the model’s ability to identify positive instances correctly.
- *Specificity:* Given by 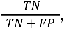 specificity quantifies the model’s ability to correctly identify negative instances.
- *Area Under the Curve (AUC):* The area under the receiver operating characteristic (ROC) curve was computed using the roc_auc_score function from scikit-learn. This metric provides a single scalar value to summarize the trade-off between sensitivity and specificity across different classification thresholds.
- *Precision:* Defined as 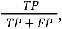 the precision metric measures the proportion of correctly predicted positive instances out of all predicted positives. This metric is particularly important in imbalanced datasets to ensure that the model does not generate a high number of false positives.
- *F1 Score*: The F1 score is the harmonic mean of precision and recall, calculated as 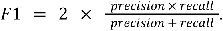
- *Matthews Correlation Coefficient (MCC)*: MCC is a balanced metric defined as 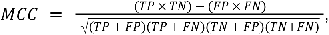 producing a score between -1 and +1, where +1 indicates perfect prediction, 0 represents no better than random guessing, and -1 signifies complete disagreement between predictions and actual values.

We used a stratified 10-fold cross-validation strategy to ensure robust performance evaluation. All classifier evaluations were conducted within individual datasets (intra-dataset evaluations), without performing cross-database evaluations.

### Ranking speech representations using a multicriteria approach

To identify the optimal speech representation for generating the best classification performance across different speech modes, we employed a Multi-Criteria Decision Analysis (MCDA). This method focuses on ranking alternatives based on multiple criteria simultaneously. By evaluating alternatives across several metrics derived from machine learning models, MCDA provided a structured framework for comparing and prioritizing options. We adopted the Technique for Order of Preference by Similarity to Ideal Solution (TOPSIS), a multicriteria approach, for ranking machine learning classifiers as outlined in [28].

#### TOPSIS MCDA

TOPSIS was selected as the MCDA method due to its alignment with machine learning tasks and its ability to incorporate the Euclidean distance from an ideal solution. In biomedical engineering, TOPSIS is frequently applied for evaluating alternatives in healthcare technology assessment, such as selecting the most appropriate medical devices [29]. For each speech mode, six speech representations were evaluated using the TOPSIS method, with each representation characterized by seven metrics—accuracy, sensitivity, specificity, AUC, precision, F1 score, and MCC—representing the average performance across multiple classifiers. TOPSIS identified the optimal alternative by minimizing the Euclidean distance to the positive ideal solution (PIS) while maximizing the distance from the negative ideal solution (NIS).

To implement TOPSIS, we first normalized the performance values of each alternative to a scale between 0 and 1 and assigned equal weights to all metrics. These weighted values were then used to compute the PIS, representing the best possible performance, and the NIS, representing the worst possible performance. The Euclidean distances from each alternative to the PIS and NIS were calculated, and alternatives were ranked based on their proximity to the PIS. Additional details regarding the implementation can be found in Supplementary Methods 2. In this study, the runtime of each speech representation was not included as a criterion for TOPSIS, as it heavily depends on hardware configurations.

## Results

### Ranking the speech representations using the multicriteria approach

The TOPSIS method was applied to compare the performance of Wav2Vec 1.0, Wav2Vec 2.0, and baseline features in HC vs. PD for classification tasks, ranking the representations across all considered speech modes. According to the TOPSIS results, the Wav2Vec 2.0 was identified as the best-performing approach overall. Specifically, the first transformer layer of Wav2Vec 2.0 demonstrated the highest performance in context-dependent tasks such as reading text and monologues across datasets, while its feature extractor proved most effective for vowel-based tasks.

Wav2Vec 1.0 ranked second among the representations, with its feature extractor performing better for vowel tasks and its combination of feature extractor and aggregator showing improved results for context-dependent modes. An exception to these general trends was observed in the dialogue speech mode within the English dataset, which showed significantly different behavior across the representations, when baselines performed the best. The TOPSIS scoring results are summarized in Table 2, while a comprehensive TOPSIS table, including detailed metrics, is provided in Supplementary Table 1.

**Table 2.** MCDA TOPSIS ranking. TOPSIS incorporated all metrics across classifiers—accuracy, sensitivity, specificity, AUC, precision, F1 score, and MCC. Green highlights indicate detected trends across datasets and modes involving context-dependent tasks, while blue highlights mark trends specific to the simpler vowel-based tasks.

| dataset | mode | rank (score) |  |  |  |  |  |
| --- | --- | --- | --- | --- | --- | --- | --- |
|  |  | 1 | 2 | 3 | 4 | 5 | 6 |
| English [17] | read text | w2v2-FT (0.88) | w2v1-FEA (0.85) | baselines (0.51) | w2v1-FE (0.41) | w2v2-FE (0.18) | w2v2-LH (0.07) |
|  | dialogue | baselines (0.95) | w2v2-FT (0.81) | w2v1-FEA (0.80) | w2v1-FE (0.66) | w2v2-FE (0.43) | w2v2-LH (0.02) |
| PC-GITA [18] | read text | w2v2-FT (0.96) | w2v1-FEA (0.89) | w2v1-FE (0.81) | w2v2-FE (0.68) | baselines (0.20) | w2v2-LH (0.10) |
|  | monologue | w2v2-FT (0.94) | w2v1-FEA (0.93) | w2v2-FE (0.87) | w2v1-FE (0.77) | w2v2-LH (0.27) | baselines (0.13) |
|  | vowels | w2v2-FE (0.95) | w2v2-FT (0.93) | w2v1-FE (0.75) | w2v1-FEA (0.44) | w2v2-LH (0.41) | baselines (0.05) |
| [19] | vowels | w2v2-FE (0.89) | w2v2-FT (0.78) | w2v1-FE (0.40) | baselines (0.30) | w2v1-FEA (0.30) | w2v2-LH (0.20) |

w2v2-FT: first transformer layer; w2v2-LH: last hidden layer; w2v2-FE: feature extraction; w2v1-FE: feature extraction; w2v1-FEA: feature extraction and aggregation

### Runtime and PCA

The Wav2Vec 1.0 was the fastest in terms of computation time, highlighting its potential for time-sensitive applications (Figure 1).

**Figure 1.**
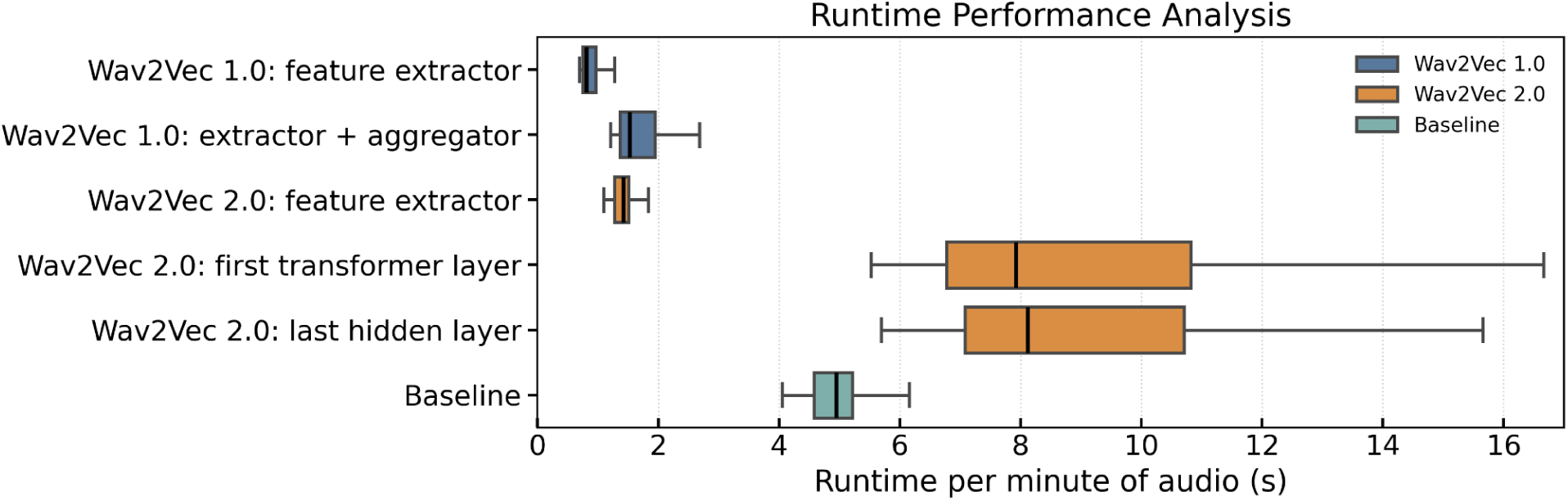
Boxplot of runtime per audio recording, normalized to one minute of audio for fair comparison across different sample lengths. Outliers were removed, including a Wav2Vec 2.0 instance exceeding 200 seconds. All measurements were performed on a MacBook Air with an M1 chip and 8GB of RAM.

For classification purposes, we computed the first 30 PCA components for all Wav2Vec representations. Figure 2 illustrates the cumulative explained variance ratio as a function of the number of PCA components.

**Figure 2.**
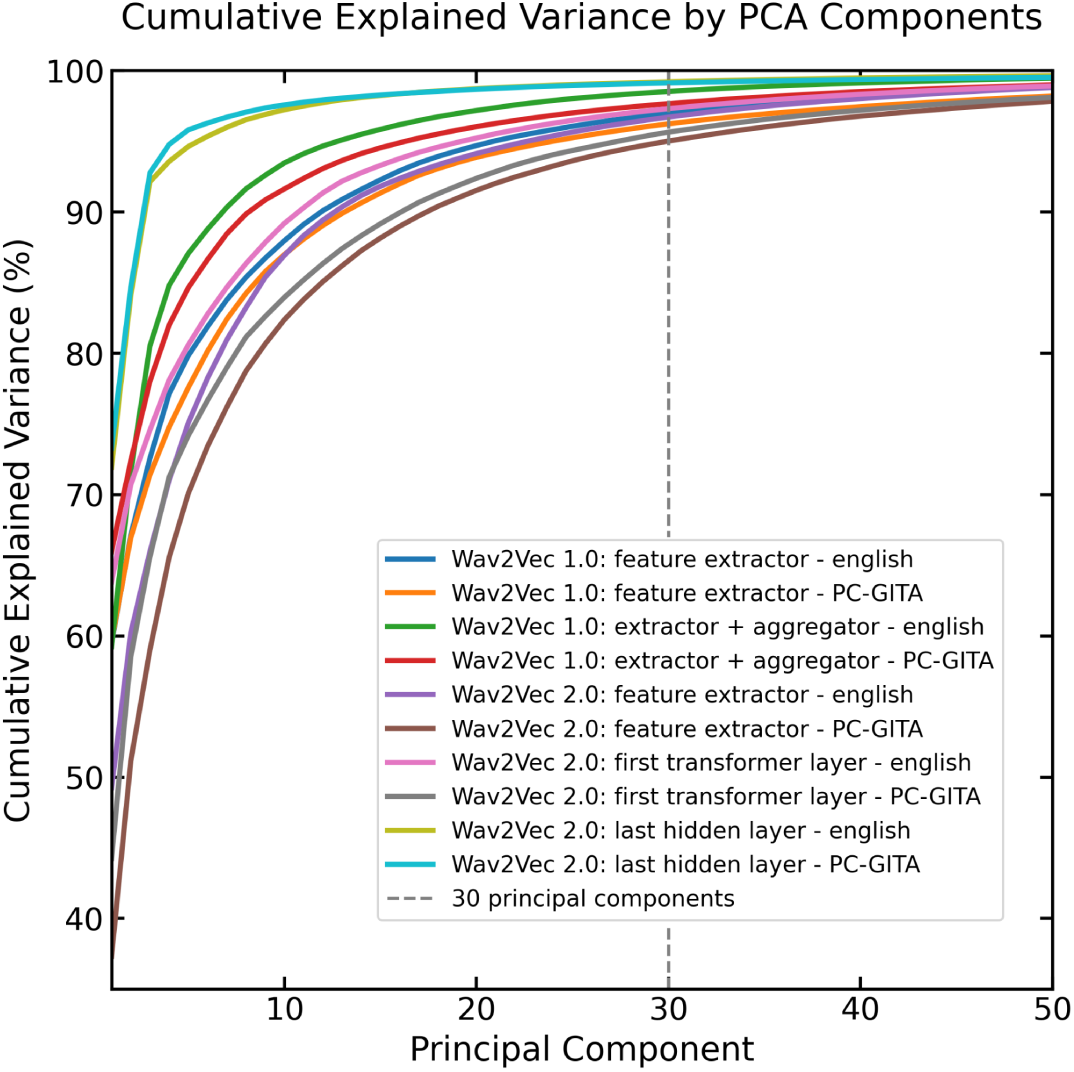
Plot of the cumulative explained variance as a function of the number of PCA components. The first 30 components captured approximately 95% of the original variance across all datasets. For Wav2Vec 2.0 representations on the PC-GITA dataset, the minimum explained variance with 30 components was 94%.

### Developed classification models across two speech modes using the English dataset

The performance of different classifiers was evaluated across two speech modes, read text and spontaneous dialogue, using the English dataset. For the read text mode, the best results across multiple metrics were achieved using the first transformer layer of the Wav2Vec 2.0 and a feature extractor and aggregator approach based on the Wav2Vec 1.0. However, there were performance variations observed across individual layers of the Wav2Vec 2.0, with the last hidden layer showing the weakest performance. Sensitivity remained low across all representations. Detailed classifier performance metrics are presented in Table 3. For spontaneous dialogue (Table 4), as a prominent result, traditional baseline features demonstrated the best overall performance, outperforming both Wav2Vec representations.

**Table 3.**
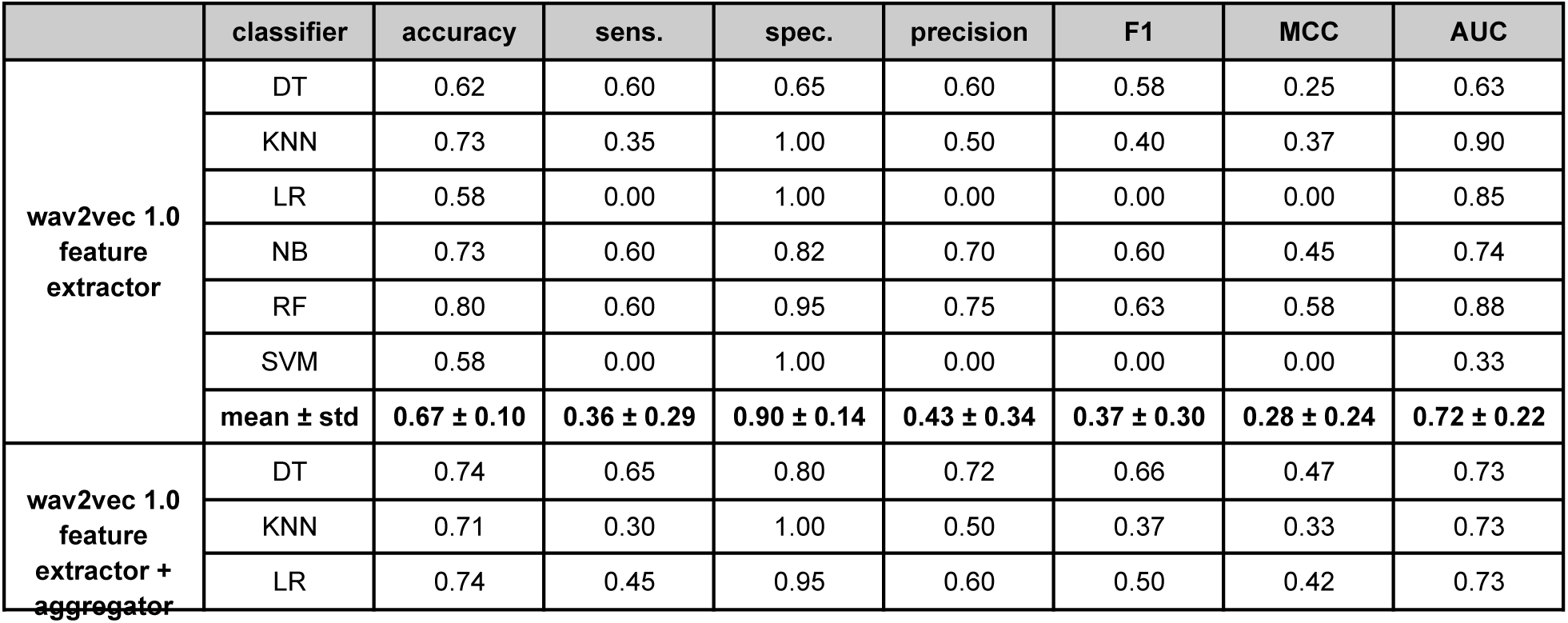
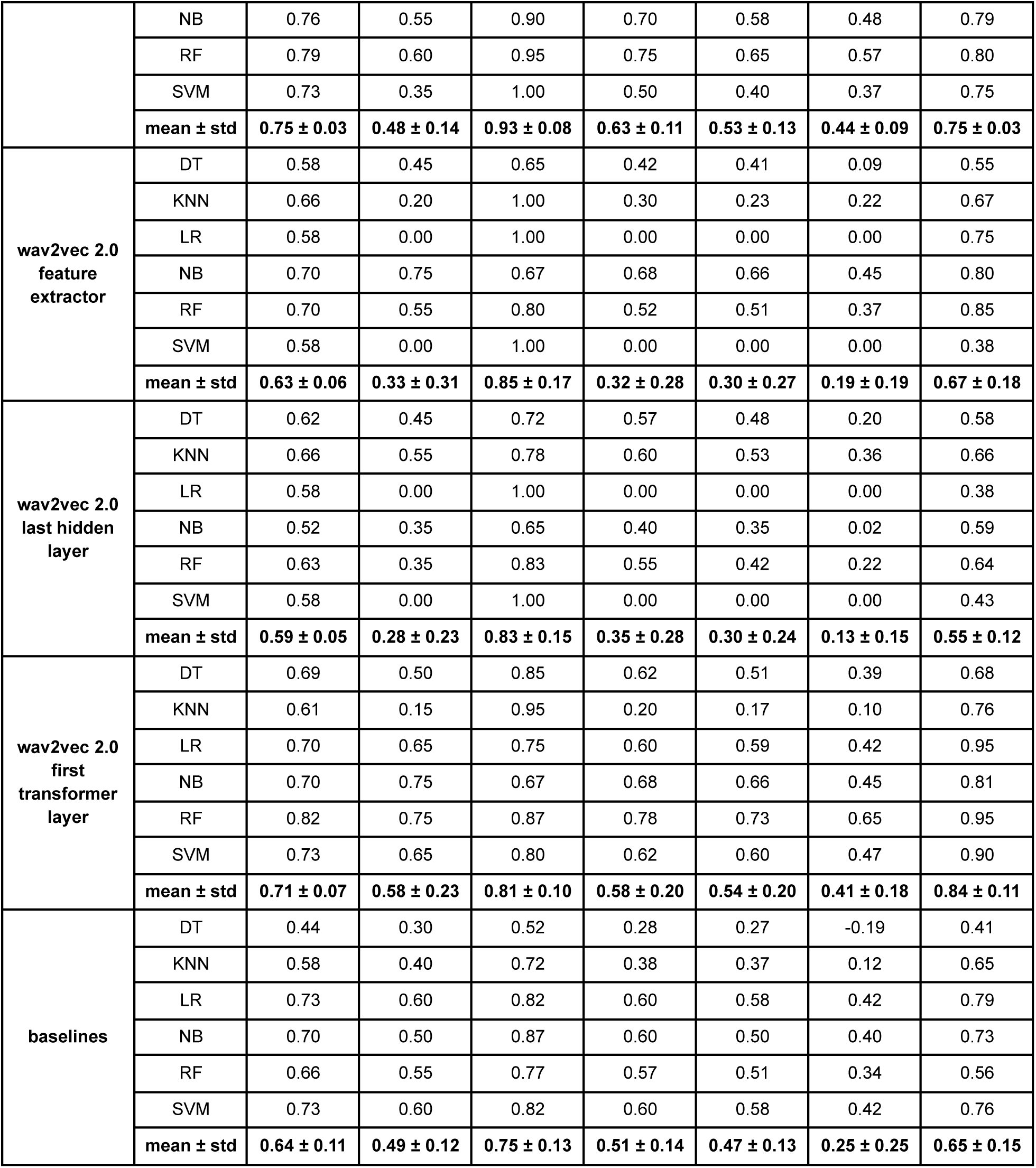
Classifier performance metrics for read text from the English dataset, which included 21 HC and 16 PD samples in an imbalanced distribution.

|  | classifier | accuracy | sens. | spec. | precision | F1 | MCC | AUC |
| --- | --- | --- | --- | --- | --- | --- | --- | --- |
| wav2vec 1.0<br>feature<br>extractor | DT | 0.62 | 0.60 | 0.65 | 0.60 | 0.58 | 0.25 | 0.63 |
|  | KNN | 0.73 | 0.35 | 1.00 | 0.50 | 0.40 | 0.37 | 0.90 |
|  | LR | 0.58 | 0.00 | 1.00 | 0.00 | 0.00 | 0.00 | 0.85 |
|  | NB | 0.73 | 0.60 | 0.82 | 0.70 | 0.60 | 0.45 | 0.74 |
|  | RF | 0.80 | 0.60 | 0.95 | 0.75 | 0.63 | 0.58 | 0.88 |
|  | SVM | 0.58 | 0.00 | 1.00 | 0.00 | 0.00 | 0.00 | 0.33 |
| | mean ± std | 0.67 ± 0.10 | 0.36 ± 0.29 | 0.90 ± 0.14 | 0.43 ± 0.34 | 0.37 ± 0.30 | 0.28 ± 0.24 | 0.72 ± 0.22 |
| wav2vec 1.0<br>feature<br>extractor +<br>aggregator | DT | 0.74 | 0.65 | 0.80 | 0.72 | 0.66 | 0.47 | 0.73 |
|  | KNN | 0.71 | 0.30 | 1.00 | 0.50 | 0.37 | 0.33 | 0.73 |
|  | LR | 0.74 | 0.45 | 0.95 | 0.60 | 0.50 | 0.42 | 0.73 |
|  | NB | 0.76 | 0.55 | 0.90 | 0.70 | 0.58 | 0.48 | 0.79 |
|  | RF | 0.79 | 0.60 | 0.95 | 0.75 | 0.65 | 0.57 | 0.80 |
|  | SVM | 0.73 | 0.35 | 1.00 | 0.50 | 0.40 | 0.37 | 0.75 |
| | mean ± std | **0.75 ± 0.03** | **0.48 ± 0.14** | **0.93 ± 0.08** | **0.63 ± 0.11** | **0.53 ± 0.13** | **0.44 ± 0.09** | **0.75 ± 0.03** |
| wav2vec 2.0<br>feature<br>extractor | DT | 0.58 | 0.45 | 0.65 | 0.42 | 0.41 | 0.09 | 0.55 |
|  | KNN | 0.66 | 0.20 | 1.00 | 0.30 | 0.23 | 0.22 | 0.67 |
|  | LR | 0.58 | 0.00 | 1.00 | 0.00 | 0.00 | 0.00 | 0.75 |
|  | NB | 0.70 | 0.75 | 0.67 | 0.68 | 0.66 | 0.45 | 0.80 |
|  | RF | 0.70 | 0.55 | 0.80 | 0.52 | 0.51 | 0.37 | 0.85 |
|  | SVM | 0.58 | 0.00 | 1.00 | 0.00 | 0.00 | 0.00 | 0.38 |
| | mean ± std | **0.63 ± 0.06** | **0.33 ± 0.31** | **0.85 ± 0.17** | **0.32 ± 0.28** | **0.30 ± 0.27** | **0.19 ± 0.19** | **0.67 ± 0.18** |
| wav2vec 2.0<br>last hidden<br>layer | DT | 0.62 | 0.45 | 0.72 | 0.57 | 0.48 | 0.20 | 0.58 |
|  | KNN | 0.66 | 0.55 | 0.78 | 0.60 | 0.53 | 0.36 | 0.66 |
|  | LR | 0.58 | 0.00 | 1.00 | 0.00 | 0.00 | 0.00 | 0.38 |
|  | NB | 0.52 | 0.35 | 0.65 | 0.40 | 0.35 | 0.02 | 0.59 |
|  | RF | 0.63 | 0.35 | 0.83 | 0.55 | 0.42 | 0.22 | 0.64 |
|  | SVM | 0.58 | 0.00 | 1.00 | 0.00 | 0.00 | 0.00 | 0.43 |
| | mean ± std | **0.59 ± 0.05** | **0.28 ± 0.23** | **0.83 ± 0.15** | **0.35 ± 0.28** | **0.30 ± 0.24** | **0.13 ± 0.15** | **0.55 ± 0.12** |
| wav2vec 2.0<br>first<br>transformer<br>layer | DT | 0.69 | 0.50 | 0.85 | 0.62 | 0.51 | 0.39 | 0.68 |
|  | KNN | 0.61 | 0.15 | 0.95 | 0.20 | 0.17 | 0.10 | 0.76 |
|  | LR | 0.70 | 0.65 | 0.75 | 0.60 | 0.59 | 0.42 | 0.95 |
|  | NB | 0.70 | 0.75 | 0.67 | 0.68 | 0.66 | 0.45 | 0.81 |
|  | RF | 0.82 | 0.75 | 0.87 | 0.78 | 0.73 | 0.65 | 0.95 |
|  | SVM | 0.73 | 0.65 | 0.80 | 0.62 | 0.60 | 0.47 | 0.90 |
| | mean ± std | **0.71 ± 0.07** | **0.58 ± 0.23** | **0.81 ± 0.10** | **0.58 ± 0.20** | **0.54 ± 0.20** | **0.41 ± 0.18** | **0.84 ± 0.11** |
| baselines | DT | 0.44 | 0.30 | 0.52 | 0.28 | 0.27 | -0.19 | 0.41 |
|  | KNN | 0.58 | 0.40 | 0.72 | 0.38 | 0.37 | 0.12 | 0.65 |
|  | LR | 0.73 | 0.60 | 0.82 | 0.60 | 0.58 | 0.42 | 0.79 |
|  | NB | 0.70 | 0.50 | 0.87 | 0.60 | 0.50 | 0.40 | 0.73 |
|  | RF | 0.66 | 0.55 | 0.77 | 0.57 | 0.51 | 0.34 | 0.56 |
|  | SVM | 0.73 | 0.60 | 0.82 | 0.60 | 0.58 | 0.42 | 0.76 |
| | mean ± std | **0.64 ± 0.11** | **0.49 ± 0.12** | **0.75 ± 0.13** | **0.51 ± 0.14** | **0.47 ± 0.13** | **0.25 ± 0.25** | **0.65 ± 0.15** |

**Table 4.** Classifier performance metrics for dialogue from the English dataset, which included 21 HC and 15 PD samples in an imbalanced distribution.

|  | classifier | accuracy | sens. | spec. | precision | F1 | MCC | AUC |
| --- | --- | --- | --- | --- | --- | --- | --- | --- |
| wav2vec 1.0<br>feature<br>extractor | DT | 0.72 | 0.75 | 0.70 | 0.65 | 0.66 | 0.47 | 0.73 |
|  | KNN | 0.72 | 0.45 | 0.92 | 0.50 | 0.47 | 0.37 | 0.61 |
|  | LR | 0.59 | 0.00 | 1.00 | 0.00 | 0.00 | 0.00 | 0.82 |
|  | NB | 0.71 | 0.60 | 0.83 | 0.53 | 0.53 | 0.44 | 0.75 |
|  | RF | 0.77 | 0.65 | 0.88 | 0.58 | 0.60 | 0.53 | 0.75 |
|  | SVM | 0.59 | 0.00 | 1.00 | 0.00 | 0.00 | 0.00 | 0.62 |
|  | **mean ± std** | **0.68 ± 0.07** | **0.41 ± 0.33** | **0.89 ± 0.11** | **0.38 ± 0.30** | **0.38 ± 0.30** | **0.30 ± 0.24** | **0.71 ± 0.08** |
| **wav2vec 1.0**<br>**feature**<br>**extractor +**<br>**aggregator** | DT | 0.48 | 0.35 | 0.58 | 0.23 | 0.27 | -0.07 | 0.47 |
|  | KNN | 0.68 | 0.55 | 0.82 | 0.50 | 0.50 | 0.35 | 0.64 |
|  | LR | 0.72 | 0.50 | 0.92 | 0.50 | 0.48 | 0.42 | 0.79 |
|  | NB | 0.70 | 0.65 | 0.78 | 0.58 | 0.57 | 0.45 | 0.70 |
|  | RF | 0.68 | 0.55 | 0.83 | 0.48 | 0.48 | 0.38 | 0.85 |
|  | SVM | 0.72 | 0.50 | 0.92 | 0.50 | 0.48 | 0.42 | 0.69 |
|  | **mean ± std** | **0.66 ± 0.09** | **0.52 ± 0.10** | **0.81 ± 0.12** | **0.47 ± 0.12** | **0.46 ± 0.10** | **0.32 ± 0.20** | **0.69 ± 0.13** |
| **wav2vec 2.0**<br>**feature**<br>**extractor** | DT | 0.52 | 0.55 | 0.57 | 0.40 | 0.40 | 0.13 | 0.56 |
|  | KNN | 0.66 | 0.40 | 0.87 | 0.45 | 0.40 | 0.27 | 0.68 |
|  | LR | 0.59 | 0.00 | 1.00 | 0.00 | 0.00 | 0.00 | 0.72 |
|  | NB | 0.63 | 0.70 | 0.62 | 0.53 | 0.57 | 0.32 | 0.75 |
|  | RF | 0.62 | 0.50 | 0.77 | 0.38 | 0.40 | 0.27 | 0.77 |
|  | SVM | 0.59 | 0.00 | 1.00 | 0.00 | 0.00 | 0.00 | 0.42 |
|  | **mean ± std** | **0.60 ± 0.05** | **0.36 ± 0.29** | **0.80 ± 0.19** | **0.29 ± 0.23** | **0.29 ± 0.24** | **0.16 ± 0.14** | **0.65 ± 0.14** |
| **wav2vec 2.0**<br>**last hidden**<br>**layer** | DT | 0.68 | 0.60 | 0.72 | 0.53 | 0.54 | 0.32 | 0.66 |
|  | KNN | 0.44 | 0.10 | 0.67 | 0.13 | 0.11 | -0.24 | 0.38 |
|  | LR | 0.59 | 0.00 | 1.00 | 0.00 | 0.00 | 0.00 | 0.33 |
|  | NB | 0.39 | 0.15 | 0.55 | 0.20 | 0.17 | -0.31 | 0.40 |
|  | RF | 0.48 | 0.20 | 0.67 | 0.25 | 0.20 | -0.13 | 0.44 |
|  | SVM | 0.59 | 0.00 | 1.00 | 0.00 | 0.00 | 0.00 | 0.53 |
|  | **mean ± std** | **0.53 ± 0.11** | **0.18 ± 0.22** | **0.77 ± 0.19** | **0.19 ± 0.20** | **0.17 ± 0.20** | **-0.06 ± 0.22** | **0.45 ± 0.12** |
| **wav2vec 2.0**<br>**first**<br>**transformer**<br>**layer** | DT | 0.57 | 0.45 | 0.67 | 0.32 | 0.36 | 0.10 | 0.56 |
|  | KNN | 0.68 | 0.55 | 0.82 | 0.53 | 0.51 | 0.37 | 0.66 |
|  | LR | 0.70 | 0.65 | 0.77 | 0.53 | 0.57 | 0.40 | 0.73 |
|  | NB | 0.67 | 0.70 | 0.67 | 0.53 | 0.57 | 0.37 | 0.83 |
|  | RF | 0.68 | 0.55 | 0.82 | 0.45 | 0.47 | 0.36 | 0.80 |
|  | SVM | 0.63 | 0.65 | 0.67 | 0.50 | 0.55 | 0.30 | 0.70 |
|  | **mean ± std** | **0.65 ± 0.05** | **0.59 ± 0.09** | **0.73 ± 0.08** | **0.48 ± 0.09** | **0.50 ± 0.08** | **0.31 ± 0.11** | **0.71 ± 0.10** |
| **baselines** | DT | 0.62 | 0.50 | 0.73 | 0.45 | 0.45 | 0.25 | 0.62 |
|  | KNN | 0.58 | 0.45 | 0.68 | 0.42 | 0.40 | 0.17 | 0.68 |
|  | LR | 0.78 | 0.70 | 0.88 | 0.78 | 0.68 | 0.61 | 0.83 |
|  | NB | 0.72 | 0.60 | 0.82 | 0.73 | 0.62 | 0.45 | 0.81 |
|  | RF | 0.66 | 0.50 | 0.78 | 0.53 | 0.48 | 0.31 | 0.63 |
|  | SVM | 0.76 | 0.65 | 0.88 | 0.68 | 0.62 | 0.56 | 0.83 |
| | mean ± std | 0.69 ± 0.08 | 0.57 ± 0.10 | 0.80 ± 0.08 | 0.60 ± 0.15 | 0.54 ± 0.11 | 0.39 ± 0.18 | 0.73 ± 0.10 |

When comparing Wav2Vec-based approaches across non-spontaneous and spontaneous speech modes, clear differences in performance were observed. For non-spontaneous speech, the Wav2Vec 1.0 FEA achieved an accuracy of 0.75 and an AUC of 0.75. In contrast, for spontaneous dialogue, its performance decreased to an accuracy of 0.66 and an AUC of 0.69. Similarly, the first transformer layer of Wav2Vec 2.0 yielded an accuracy of 0.71 and an AUC of 0.84 for non-spontaneous speech, while its performance dropped to an accuracy of 0.65 and an AUC of 0.71 for spontaneous dialogue. These results highlight a consistent trend of higher performance in non-spontaneous speech compared to spontaneous dialogue, with differences in accuracy ranging from 6% to 9% and differences in the AUC ranging from 4% to 13%, depending on the representation.

Performance variability was observed across both representations and classifiers throughout the English dataset. Importantly, logistic regression and SVM classifiers failed in several instances, misclassifying all test examples as HC, resulting in a sensitivity of 0 and specificity of 1. Supplementary Tables 2 and 3 provide a comparison of PCA-reduced Wav2Vec embeddings for both speech modes.

### Developed classification models across two speech modes using the PC-GITA dataset

The performance of different classifiers was evaluated across read text (Table 5) and spontaneous monologue (Table 6) using the PC-GITA dataset. The first transformer layer of Wav2Vec 2.0 delivered the best overall performance for both speech modes, consistently outperforming other representations. Within Wav2Vec 1.0, the FEA combination significantly enhanced performance compared to the feature extractor FE alone, making it competitive with the top-performing representations.

**Table 5.** Classifier performance metrics for read text from the PC-GITA dataset, which included 50 HC and 50 PD samples in a balanced distribution.

|  | classifier | accuracy | sens. | spec. | precision | F1 | MCC | AUC |
| --- | --- | --- | --- | --- | --- | --- | --- | --- |
| wav2vec 1.0<br>feature<br>extractor | DT | 0.75 | 0.84 | 0.66 | 0.74 | 0.77 | 0.54 | 0.75 |
|  | KNN | 0.75 | 0.60 | 0.90 | 0.88 | 0.69 | 0.54 | 0.83 |
|  | LR | 0.69 | 0.58 | 0.80 | 0.76 | 0.64 | 0.40 | 0.75 |
|  | NB | 0.74 | 0.62 | 0.86 | 0.84 | 0.70 | 0.51 | 0.79 |
|  | RF | 0.85 | 0.82 | 0.88 | 0.89 | 0.84 | 0.72 | 0.90 |
|  | SVM | 0.66 | 0.50 | 0.82 | 0.75 | 0.59 | 0.35 | 0.34 |
| | mean ± std | 0.74 ± 0.07 | 0.66 ± 0.14 | 0.82 ± 0.09 | 0.81 ± 0.07 | 0.70 ± 0.09 | 0.51 ± 0.13 | 0.73 ± 0.20 |
| wav2vec 1.0<br>feature<br>extractor +<br>aggregator | DT | 0.73 | 0.78 | 0.68 | 0.72 | 0.74 | 0.47 | 0.73 |
|  | KNN | 0.75 | 0.60 | 0.90 | 0.88 | 0.69 | 0.54 | 0.81 |
|  | LR | 0.75 | 0.68 | 0.82 | 0.80 | 0.71 | 0.52 | 0.87 |
|  | NB | 0.71 | 0.64 | 0.78 | 0.75 | 0.67 | 0.44 | 0.79 |
|  | RF | 0.78 | 0.78 | 0.78 | 0.79 | 0.78 | 0.57 | 0.87 |
|  | SVM | 0.81 | 0.76 | 0.86 | 0.86 | 0.78 | 0.65 | 0.90 |
| | mean ± std | 0.76 ± 0.04 | 0.71 ± 0.08 | 0.80 ± 0.08 | 0.80 ± 0.06 | 0.73 ± 0.04 | 0.53 ± 0.07 | 0.83 ± 0.06 |
| wav2vec 2.0<br>feature<br>extractor | DT | 0.57 | 0.56 | 0.58 | 0.59 | 0.56 | 0.15 | 0.57 |
|  | KNN | 0.75 | 0.66 | 0.84 | 0.83 | 0.71 | 0.53 | 0.83 |
|  | LR | 0.68 | 0.76 | 0.60 | 0.70 | 0.71 | 0.37 | 0.77 |
|  | NB | 0.68 | 0.76 | 0.60 | 0.69 | 0.71 | 0.37 | 0.73 |
|  | RF | 0.82 | 0.80 | 0.84 | 0.88 | 0.81 | 0.67 | 0.89 |
|  | SVM | 0.69 | 0.78 | 0.60 | 0.71 | 0.73 | 0.39 | 0.76 |
| | mean ± std | **0.70 ± 0.08** | **0.72 ± 0.09** | **0.68 ± 0.13** | **0.73 ± 0.10** | **0.70 ± 0.08** | **0.42 ± 0.17** | **0.76 ± 0.11** |
| wav2vec 2.0<br>last hidden<br>layer | DT | 0.58 | 0.56 | 0.60 | 0.54 | 0.54 | 0.15 | 0.58 |
|  | KNN | 0.56 | 0.48 | 0.64 | 0.58 | 0.51 | 0.13 | 0.62 |
|  | LR | 0.40 | 0.32 | 0.48 | 0.39 | 0.34 | -0.21 | 0.38 |
|  | NB | 0.58 | 0.44 | 0.72 | 0.60 | 0.49 | 0.17 | 0.60 |
|  | RF | 0.73 | 0.76 | 0.70 | 0.76 | 0.74 | 0.48 | 0.80 |
|  | SVM | 0.45 | 0.18 | 0.72 | 0.33 | 0.21 | -0.12 | 0.56 |
| | mean ± std | **0.55 ± 0.12** | **0.46 ± 0.20** | **0.64 ± 0.09** | **0.53 ± 0.15** | **0.47 ± 0.18** | **0.10 ± 0.25** | **0.59 ± 0.13** |
| wav2vec 2.0<br>first<br>transformer<br>layer | DT | 0.78 | 0.80 | 0.76 | 0.78 | 0.78 | 0.58 | 0.78 |
|  | KNN | 0.76 | 0.68 | 0.84 | 0.84 | 0.73 | 0.55 | 0.78 |
|  | LR | 0.82 | 0.80 | 0.84 | 0.86 | 0.82 | 0.66 | 0.89 |
|  | NB | 0.68 | 0.80 | 0.56 | 0.67 | 0.72 | 0.37 | 0.74 |
|  | RF | 0.79 | 0.78 | 0.80 | 0.85 | 0.78 | 0.61 | 0.86 |
|  | SVM | 0.82 | 0.80 | 0.84 | 0.86 | 0.81 | 0.66 | 0.88 |
| | mean ± std | **0.78 ± 0.05** | **0.78 ± 0.05** | **0.77 ± 0.11** | **0.81 ± 0.07** | **0.77 ± 0.04** | **0.57 ± 0.11** | **0.82 ± 0.06** |
| baselines | DT | 0.55 | 0.62 | 0.48 | 0.54 | 0.57 | 0.10 | 0.55 |
|  | KNN | 0.57 | 0.60 | 0.54 | 0.55 | 0.57 | 0.14 | 0.61 |
|  | LR | 0.60 | 0.62 | 0.58 | 0.61 | 0.59 | 0.20 | 0.63 |
|  | NB | 0.51 | 0.66 | 0.36 | 0.49 | 0.56 | 0.05 | 0.50 |
|  | RF | 0.61 | 0.62 | 0.60 | 0.58 | 0.58 | 0.23 | 0.58 |
|  | SVM | 0.60 | 0.60 | 0.60 | 0.62 | 0.59 | 0.21 | 0.65 |
| | mean ± std | **0.57 ± 0.04** | **0.62 ± 0.02** | **0.53 ± 0.09** | **0.57 ± 0.05** | **0.58 ± 0.01** | **0.15 ± 0.07** | **0.59 ± 0.05** |

**Table 6.**
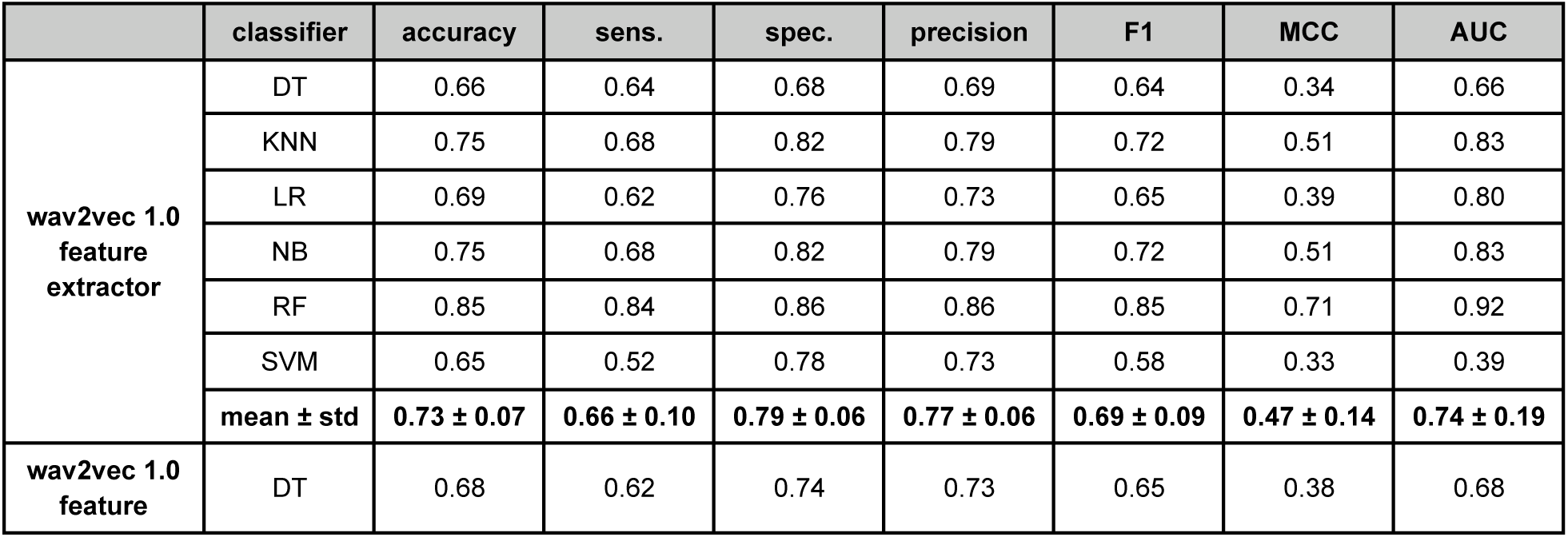
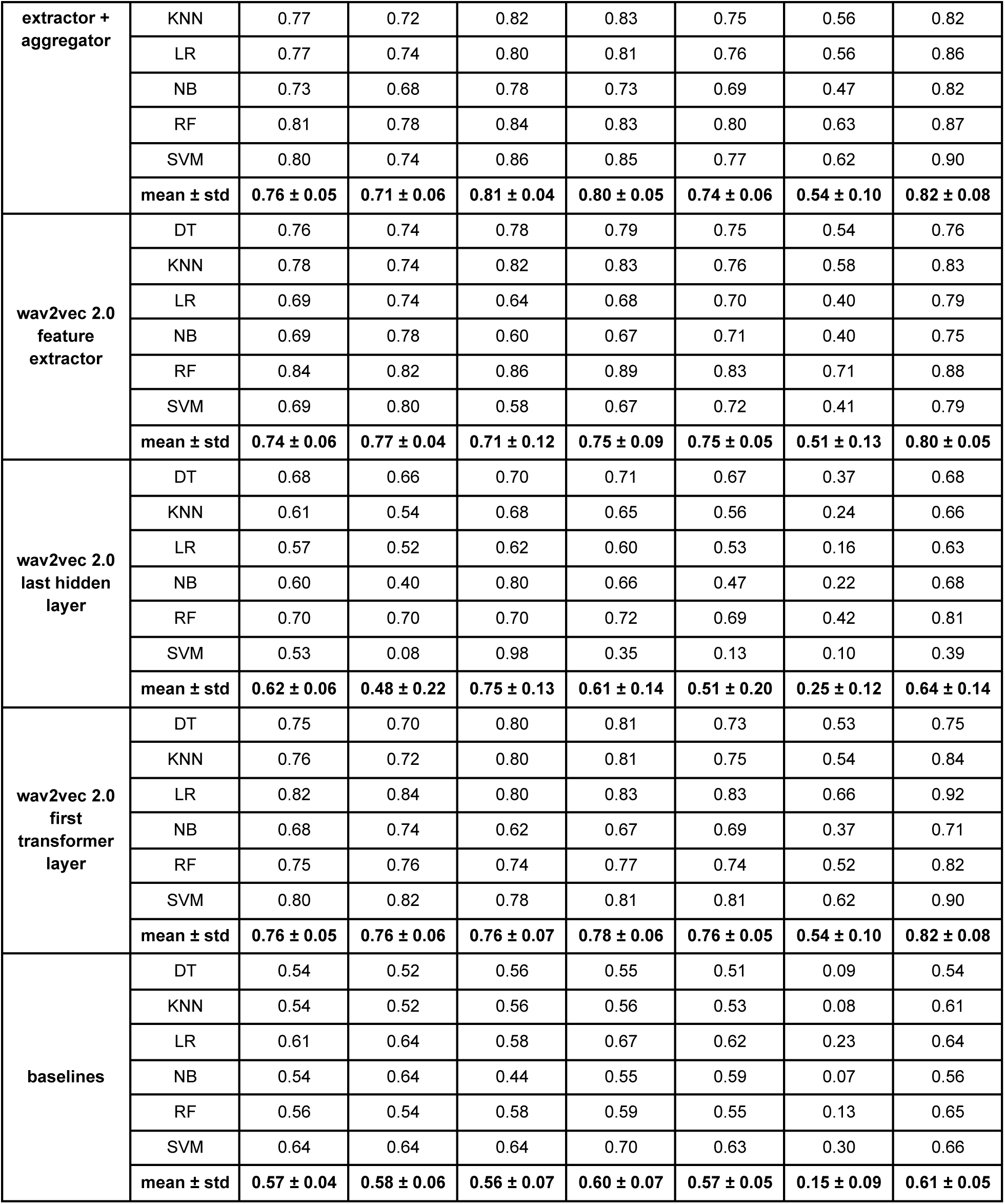
Classifier performance metrics for monologue from the PC-GITA dataset, which included 50 HC and 50 PD samples in a balanced distribution.

For both read text and monologue, the best performing Wav2Vec-based approaches demonstrated comparable results, with no significant performance differences observed between the two speech modes. However, the last hidden state of Wav2Vec 2.0 was the poorest-performing representation. Baseline features underperformed compared to Wav2Vec embeddings, with performance drops exceeding 20% across several metrics.

In contrast to the English dataset, results from PC-GITA exhibited lower variability across classifiers and more stable outcomes for higher-performing Wav2Vec representations. Further details on PCA-reduced Wav2Vec embeddings for the PC-GITA dataset are provided in Supplementary Tables 4 and 5.

### Developed classification models using the sustained vowel /a/

We further evaluated the performance of HC vs. PD classifiers on two vowel datasets, one from PC-GITA (Table 7) and the other from a U.S.-based dataset (Table 8) [19]. The feature extractor of Wav2Vec 2.0 consistently achieved the best performance across both datasets. Wav2Vec 2.0’s feature extractor yielded an accuracy of 0.69 and an AUC of 0.76 for the PC-GITA dataset, and an accuracy of 0.70 with an AUC of 0.75 for the US-based dataset. In addition, the feature extractor of Wav2Vec 1.0 outperformed its feature extractor and aggregator combination, suggesting that non-contextual configurations of the Wav2Vec 1.0 might be better suited for vowel classification tasks. Further comparisons, including PCA-reduced Wav2Vec embeddings for the vowel datasets, are detailed in Supplementary Tables 6 and 7.

**Table 7.**
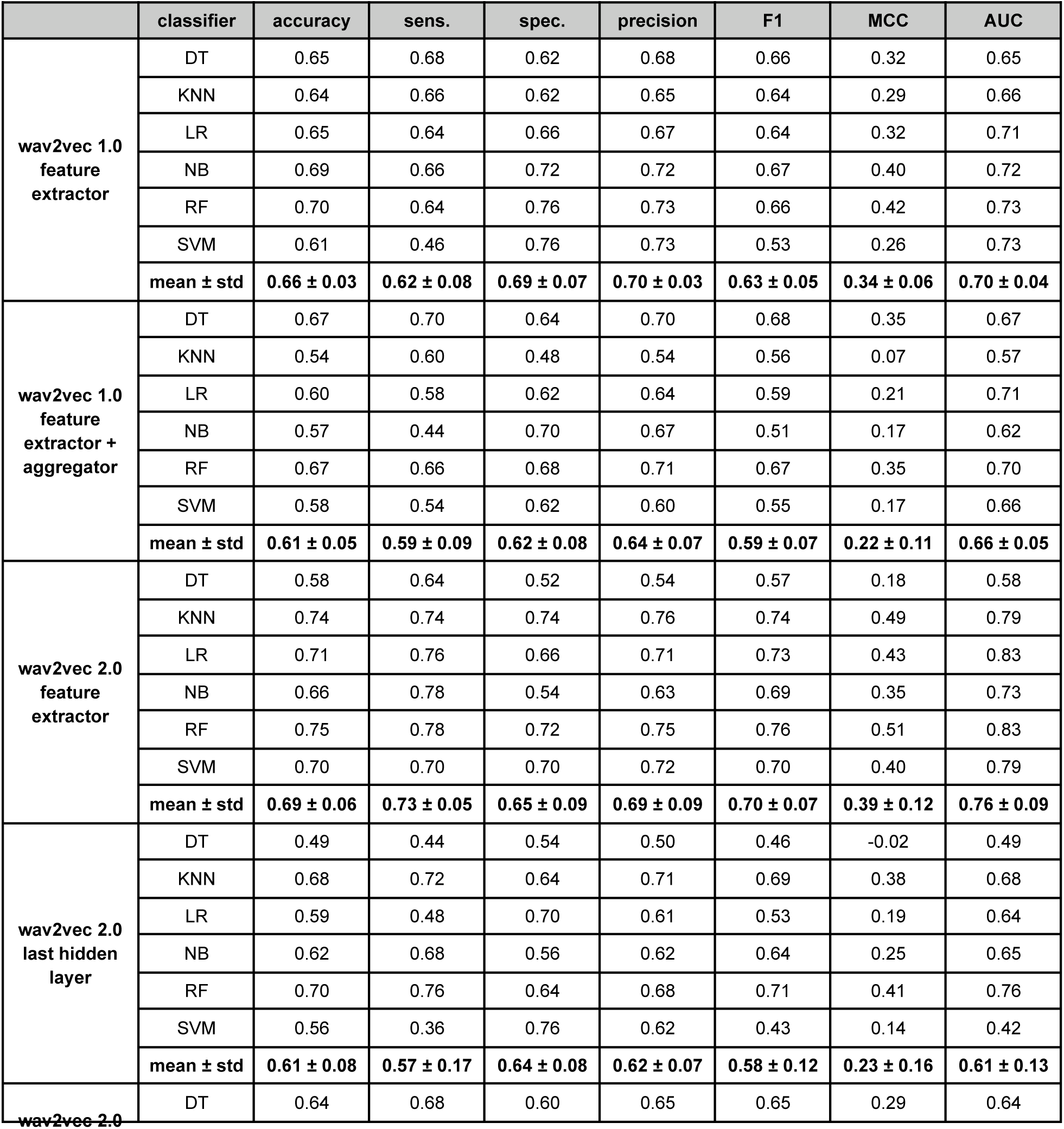
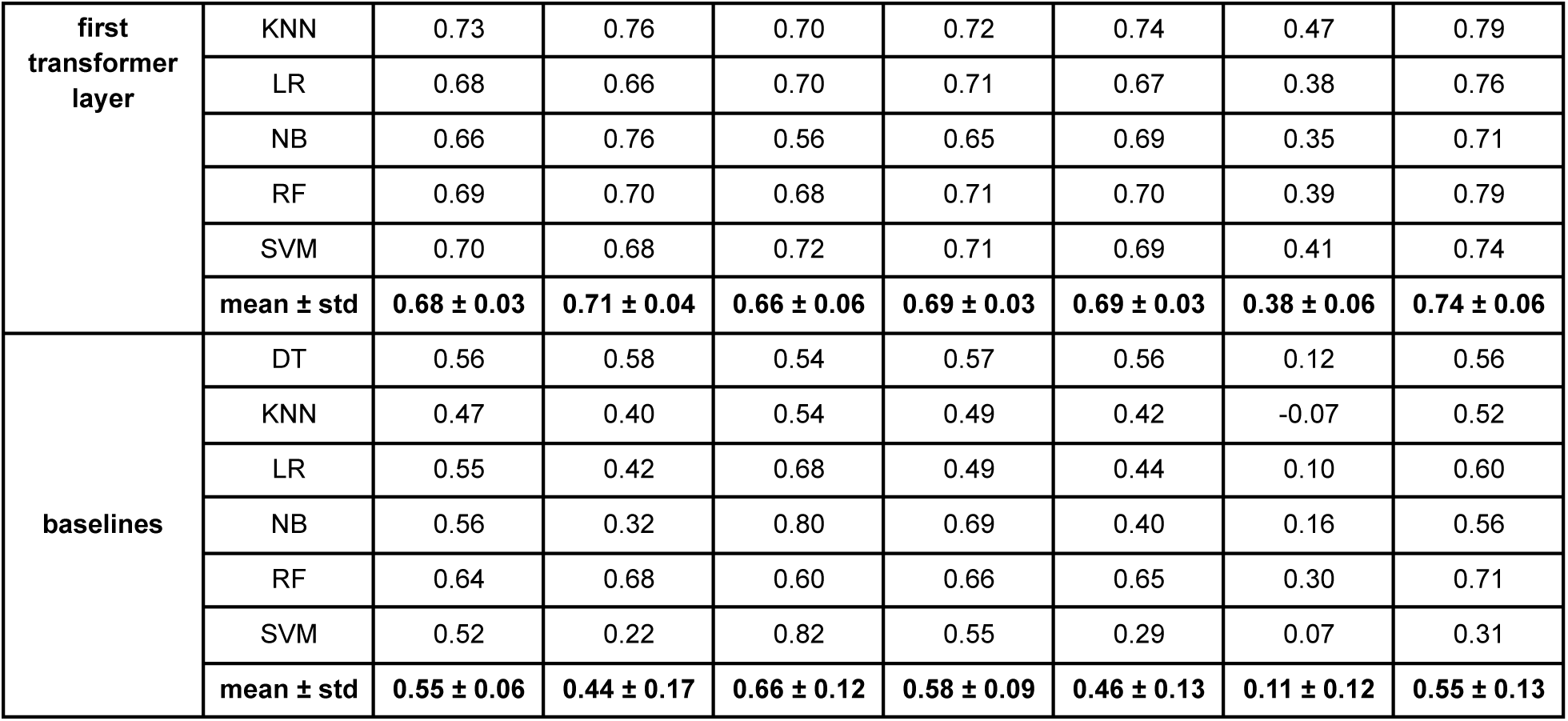
Classifier performance metrics for vowel dataset from the PC-GITA dataset, which included 50 HC and 50 PD samples in a balanced distribution.

**Table 8.** Classifier performance metrics for vowel dataset from the U.S.-based dataset, which included 41 HC and 40 PD samples in a balanced distribution.

|  | classifier | accuracy | sens. | spec. | precision | F1 | MCC | AUC |
| --- | --- | --- | --- | --- | --- | --- | --- | --- |
| wav2vec 1.0 feature extractor | DT | 0.65 | 0.70 | 0.61 | 0.64 | 0.66 | 0.32 | 0.65 |
|  | KNN | 0.54 | 0.68 | 0.42 | 0.58 | 0.58 | 0.11 | 0.62 |
|  | LR | 0.63 | 0.65 | 0.62 | 0.64 | 0.63 | 0.28 | 0.68 |
|  | NB | 0.61 | 0.50 | 0.71 | 0.62 | 0.54 | 0.22 | 0.64 |
|  | RF | 0.69 | 0.65 | 0.74 | 0.75 | 0.66 | 0.42 | 0.77 |
|  | SVM | 0.63 | 0.63 | 0.64 | 0.66 | 0.61 | 0.29 | 0.39 |
| | mean ± std | **0.63 ± 0.05** | **0.63 ± 0.07** | **0.62 ± 0.11** | **0.65 ± 0.06** | **0.61 ± 0.05** | **0.27 ± 0.10** | **0.62 ± 0.13** |
| wav2vec 1.0 feature extractor + aggregator | DT | 0.57 | 0.68 | 0.46 | 0.59 | 0.60 | 0.14 | 0.57 |
|  | KNN | 0.65 | 0.68 | 0.63 | 0.70 | 0.66 | 0.33 | 0.69 |
|  | LR | 0.62 | 0.65 | 0.59 | 0.65 | 0.62 | 0.26 | 0.65 |
|  | NB | 0.64 | 0.58 | 0.71 | 0.59 | 0.58 | 0.30 | 0.67 |
|  | RF | 0.56 | 0.55 | 0.57 | 0.50 | 0.51 | 0.13 | 0.66 |
|  | SVM | 0.62 | 0.65 | 0.58 | 0.62 | 0.62 | 0.24 | 0.69 |
| | mean ± std | **0.61 ± 0.04** | **0.63 ± 0.05** | **0.59 ± 0.08** | **0.61 ± 0.07** | **0.60 ± 0.05** | **0.23 ± 0.08** | **0.65 ± 0.05** |
| wav2vec 2.0 feature extractor | DT | 0.60 | 0.68 | 0.53 | 0.59 | 0.62 | 0.22 | 0.60 |
|  | KNN | 0.65 | 0.73 | 0.58 | 0.68 | 0.66 | 0.34 | 0.67 |
|  | LR | 0.71 | 0.75 | 0.68 | 0.72 | 0.72 | 0.45 | 0.80 |
|  | NB | 0.74 | 0.75 | 0.73 | 0.74 | 0.74 | 0.50 | 0.78 |
|  | RF | 0.75 | 0.78 | 0.73 | 0.76 | 0.76 | 0.51 | 0.82 |
|  | SVM | 0.73 | 0.75 | 0.70 | 0.74 | 0.72 | 0.48 | 0.83 |
| | mean ± std | **0.70 ± 0.06** | **0.74 ± 0.03** | **0.66 ± 0.08** | **0.70 ± 0.06** | **0.70 ± 0.05** | **0.42 ± 0.11** | **0.75 ± 0.09** |
| wav2vec 2.0<br>last hidden<br>layer | DT | 0.64 | 0.65 | 0.64 | 0.69 | 0.64 | 0.29 | 0.64 |
|  | KNN | 0.64 | 0.75 | 0.53 | 0.62 | 0.67 | 0.29 | 0.71 |
|  | LR | 0.62 | 0.40 | 0.83 | 0.59 | 0.46 | 0.24 | 0.72 |
|  | NB | 0.60 | 0.48 | 0.73 | 0.68 | 0.53 | 0.23 | 0.73 |
|  | RF | 0.65 | 0.65 | 0.65 | 0.68 | 0.63 | 0.33 | 0.78 |
|  | SVM | 0.51 | 0.00 | 1.00 | 0.00 | 0.00 | 0.00 | 0.29 |
| | mean ± std | **0.61 ± 0.06** | **0.49 ± 0.27** | **0.73 ± 0.17** | **0.54 ± 0.27** | **0.49 ± 0.25** | **0.23 ± 0.12** | **0.64 ± 0.18** |
| wav2vec 2.0<br>first<br>transformer<br>layer | DT | 0.61 | 0.58 | 0.64 | 0.58 | 0.56 | 0.22 | 0.61 |
|  | KNN | 0.67 | 0.68 | 0.66 | 0.71 | 0.65 | 0.37 | 0.72 |
|  | LR | 0.68 | 0.65 | 0.70 | 0.75 | 0.66 | 0.39 | 0.73 |
|  | NB | 0.76 | 0.75 | 0.78 | 0.78 | 0.76 | 0.54 | 0.78 |
|  | RF | 0.73 | 0.73 | 0.73 | 0.73 | 0.72 | 0.46 | 0.80 |
|  | SVM | 0.62 | 0.65 | 0.59 | 0.65 | 0.63 | 0.27 | 0.73 |
| | mean ± std | **0.68 ± 0.06** | **0.67 ± 0.06** | **0.68 ± 0.07** | **0.70 ± 0.07** | **0.66 ± 0.07** | **0.38 ± 0.12** | **0.73 ± 0.07** |
| baselines | DT | 0.54 | 0.55 | 0.53 | 0.61 | 0.53 | 0.11 | 0.54 |
|  | KNN | 0.61 | 0.50 | 0.72 | 0.68 | 0.55 | 0.24 | 0.64 |
|  | LR | 0.64 | 0.50 | 0.78 | 0.74 | 0.58 | 0.32 | 0.73 |
|  | NB | 0.63 | 0.43 | 0.83 | 0.79 | 0.53 | 0.31 | 0.67 |
|  | RF | 0.57 | 0.55 | 0.59 | 0.60 | 0.55 | 0.16 | 0.65 |
|  | SVM | 0.65 | 0.40 | 0.91 | 0.78 | 0.50 | 0.36 | 0.73 |
| | mean ± std | **0.61 ± 0.04** | **0.49 ± 0.06** | **0.73 ± 0.14** | **0.70 ± 0.09** | **0.54 ± 0.02** | **0.25 ± 0.10** | **0.66 ± 0.07** |

## Discussion

This study showed the superior performance of the Wav2Vec 2.0 across all datasets and speech modes, while also demonstrating that the Wav2Vec 1.0 remained highly competitive and, in certain cases, delivered comparable results. By employing the TOPSIS multicriteria decision analysis, we systematically ranked the different speech representations based on average performance metrics across classifiers. Our findings emphasize the importance of carefully selecting an appropriate layer when applying Wav2Vec-based approaches for PD detection, as performance might vary depending on the layer utilized. The Wav2Vec 1.0 was the fastest in terms of runtime.

An important consideration for this research, was the use of models trained on English versus multilingual datasets. Experiments with the Wav2Vec 2.0 XLSR-53, a multilingual model, and the Wav2Vec 1.0 Large, trained exclusively on English, provide insights into cross-linguistic generalizability. For instance, the competitive performance of the Wav2Vec 1.0 Large on the Spanish PC-GITA dataset, compared to the multilingual Wav2Vec 2.0 XLSR-53, suggested that classification cues distinguishing HC from PD patients are language-independent. For the vowel task, language is unlikely to play a significant role, and any observed differences are more likely attributed to the quality of the recordings.

For the English dataset, as a quantitative comparison with recent studies from 2024, Cesare et al. [30] achieved over 90% accuracy, better sensitivity and better specificity for both read text and spontaneous dialogue using non-deep learning cepstral features. While these results surpassed the classification metrics observed in this study, the AUC and other additional metrics were not reported in [30]. Additionaly, our previous study [16], focused on the read-text task using Wav2Vec 1.0 and a random forest classifier. The performance of Wav2Vec 1.0 in that study aligned with our current observations, as expected. In addition, this study offered several classifiers comparison and performance across metrics, including accuracy, sensitivity, specificity, F1 score, and MCC. Importantly, we observed an improvement of up to 15% in the AUC with the Wav2Vec 2.0 compared to the Wav2Vec 1.0.

For the PC-GITA dataset, recent studies from 2024 have identified the Wav2Vec 2.0 as a top-tier-performing foundational speech model. One such study [10] utilized the base model of the Wav2Vec 2.0 and reported an average accuracy of 80% and an AUC of 0.8, results that are consistent with the findings presented in this study. Building on this, significant improvements were demonstrated in the monologue task when frame-wise information extracted from the Wav2Vec 2.0 representations was incorporated [31]. Further advancements in PD detection include the application of diffusion-based conditional generative models for speech enhancement [32]. This method separated clean signals and residue signals from the original recordings, leading to enhanced classifier performance for dysarthric speech. Relevant to this study, the inputs for generative models were presented there as “Original signals”. When paired with the Wav2Vec 2.0 representation and multilayer perceptron classifier, the AUC achieved values of 0.83, accuracy of 0.79, sensitivity of 0.74, and specificity of 84% [32]. These findings align closely with our observations.

For the sustained vowel /a/ datasets, prior studies have demonstrated that specific algorithms and approaches can outperform wav2vec-based methods in certain aspects and wav2vec can be more suitable for complex speech tasks rather than simpler tasks like sustained vowel classification. Hires et al. achieved an exceptional performance on the PC-GITA dataset using a CNN model, reporting an AUC exceeding 0.9 [33]. Our findings align more closely with their results obtained using a comprehensive set of traditional, non-deep learning features, which achieved an average AUC of 0.88—approximately 5% higher than the best AUC observed with Random Forest classifiers in this study. On the U.S.-based vowels dataset, our Random Forest w2v2-based approach achieved an accuracy of 0.75, slightly outperforming their results across four feature sets, where the best reported accuracy was 0.73 [19]. Additionally, their research highlighted a transfer-learned CNN solution, which achieved a performance comparable to the CNN by Hires et al. Nevertheless, Wav2Vec embeddings proved to be relatively competitive with other state-of-the-art methods, offering a viable alternative for sustained vowel tasks, particularly when considering their broader applicability across diverse speech modes and disorders.

In the context of [11], our findings are highly comparable, particularly regarding PC-GITA and Wav2Vec 2.0. The referenced study reported an average accuracy of 77% for non-spontaneous speech and 75% for spontaneous speech, focusing exclusively on embeddings extracted from the first transformer layer of the XLSR-53 model. In this study, we extended these findings by analyzing outputs from additional layers of the Wav2Vec 2.0 architecture. Furthermore, as discussed in [11], the last layers of the Wav2Vec 2.0 models are better suited for tasks involving phonetic or non-paralinguistic content, such as sound production, while the first layers are more aligned with paralinguistic and prosody-related tasks, such as rhythm and intonation, often extending into contextual speech. This aligns with our observations, where the feature extractor of the Wav2Vec 2.0 performed better for vowel classification, while the first transformer layer excelled in contextual speech tasks.

Importantly, our results also demonstrated consistency with the previously reported compensatory patterns between spontaneous and non-spontaneous speech modes in Wav2Vec 2.0-based models. These models effectively extract additional cues in spontaneous speech that are less accessible in non-spontaneous speech, as highlighted in [11]. Interestingly, we observed a similar trend of compensation with the Wav2Vec 1.0, extending this behavior across both architectures. In contrast, new results in this study reveal no evidence of compensation between spontaneous dialogue and read text in the English dataset. This observation differed from the trends noted in PC-GITA and could be a possible limitation of this study, as discussed further in the next paragraph.

Although Wav2Vec-based representations generally outperformed baseline features, some exceptions were observed. For example, in the dialogue speech mode for the English dataset, baseline features achieved the best performance. This anomaly may be attributed to the interaction dynamics between the subject and the test executor, where the switching of speakers potentially disrupted the contextual cues captured by Wav2Vec representations, leading to reduced performance. Partitioning an audio stream into homogeneous segments based on speaker identity, as implemented in [30], and processing the remaining content similar to monologues in datasets like PC-GITA, could enhance classification outcomes in such cases. In our future work, this approach might also reveal compensatory patterns between non-spontaneous and spontaneous speech when using Wav2Vec-based methods.

An important aspect of our findings concerned the variability and influence of downstream classifiers. Random Forest was one of the most stable and consistently high-performing classifiers, which aligns with its ability to effectively identify key features within high-dimensional data. Some other classifiers displayed higher variability, with some showing larger standard deviations in performance, making them less reliable in this context. PCA was applied to reduce the dimensionality of the wav2vec representations. Across speech modes, classification on the English dataset using 30 PCA components resulted in a significant drop in performance. Similar or slightly reduced performance was also observed for PC-GITA and vowel datasets compared to classification using the full feature set. This indicated that while dimensionality reduction can simplify data processing, it may lead to the loss of critical information for the deep embeddings.

This study has certain limitations. The input performance metrics used for the TOPSIS rankings were highly dependent and correlated, and we did not conduct sensitivity analyses or experiment with weighting different metrics. Additionally, fine-tuning both classifiers and Wav2Vec architectures was not explored in this study. Fine-tuning the wav2vec-based classifiers specifically for HC vs. PD classification has been investigated in prior research [8]. Fine-tuning, in general, is highly computationally demanding. Despite these challenges, the Wav2Vec 2.0 remains a promising candidate for further modifications and downstream experiments involving layer selection and block combinations. Approaches such as classifier combination and attention-based feature fusion, as presented in [4], could be explored to establish optimal layer utilization strategies and achieve state-of-the-art results comparable to those containing fine-tuning. While such strategies have been applied to tasks like emotion detection, they have yet to be addressed for dysarthric speech. Notably, fine-tuning the Wav2Vec 2.0 for dysarthria has been attempted by Jamanvardi et al. [34], setting a precedent for future work in this domain.

## Conclusion

This study established the Wav2Vec 2.0 as the top-performing speech representation technique for PD detection across non-spontaneous, spontaneous, and vowel-based classifications. Its consistent superiority highlighted its utility in identifying PD-related speech characteristics. However, the Wav2Vec 1.0 should not be overlooked. Despite its simpler, non-transformer architecture, it demonstrated competitive performance and remained a valuable alternative, particularly when rapid computation is prioritized over marginal performance gains. Its faster processing times make it a promising option for applications like federated learning, where computational efficiency, data anonymization, compression, and secure distributed analysis are essential. Our findings extend existing theories on disorder classification using the Wav2Vec by offering new insights into the behavior of different speech modes. Future research should explore the potential fusion of the Wav2Vec 1.0 and 2.0 to determine whether their complementary characteristics can capture additional types of information relevant to PD detection.

## Data Availability

The datasets used in this study are publicly available, with one dataset being available for download upon request from their authors.

1. https://doi.org/10.5281/zenodo.2867216

2. https://figshare.com/articles/dataset/Voice_Samples_for_Patients_with_Parkinson_s_Disease_and_Healthy_Controls/23849127

3. The PC-GITA dataset is available upon request from Juan Rafael Orozco-Arroyave affiliated with Universidad de Antioquia UdeA.

https://doi.org/10.5281/zenodo.2867216

https://figshare.com/articles/dataset/Voice_Samples_for_Patients_with_Parkinson_s_Disease_and_Healthy_Controls/23849127

## Acknowledgement

This study was supported by the project of the National Institute for Neurological Research (Programme EXCELES, ID Project No. LX22NPO5107)—funded by the European Union—Next Generation EU.

## Author contribution

The authors confirm contribution to the paper as follows: study conception and design: OK, RK, AT; data collection: AS, OK; analysis and interpretation of results: AS, OK; draft manuscript preparation: OK. All authors reviewed the results and approved the final version of the manuscript.

