## Supplementary Materials for "Ranking Pretrained Speech Embeddings in Parkinson’s Disease Detection: Does Wav2Vec 2.0 Outperform its 1.0 Version Across Speech Modes and Languages?"

### Supplementary Figures

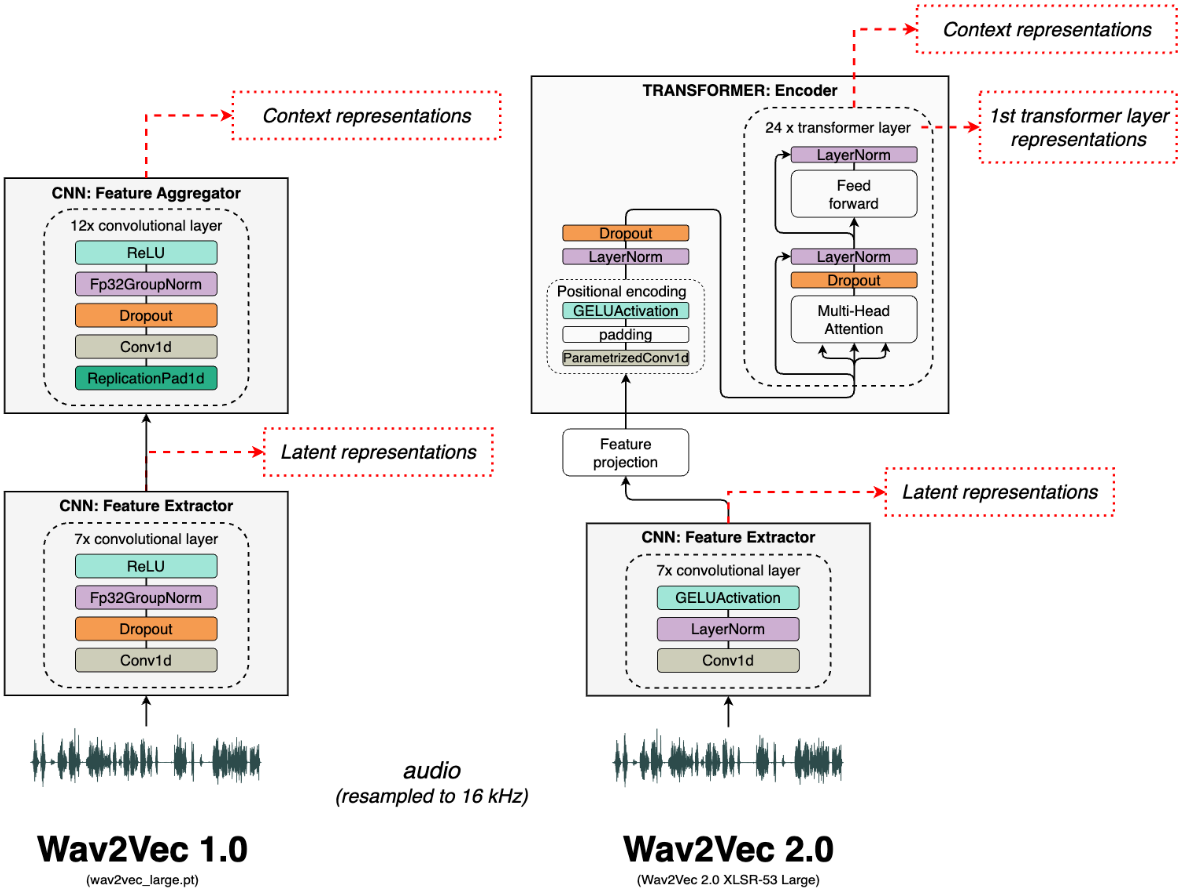


**Figure S1.** Two architecture versions of the Wav2Vec used.

### Supplementary Methods 1 - baseline features

The following audio frame features (A-F) were extracted for each audio signal:

A. Energy Entropy

$$EE_t = -\sum_{s=1}^{S} s * log_2 s, \quad (1)$$

the time domain signal for frame $t$ is split into 10 short blocks. $S$ is the number of short blocks, $s$ corresponds to the energy of a short block.

B. Energy

$$E_t = \sqrt{\frac{1}{N}\sum_{n=1}^{N} |x[n]|^2}, \tag{2}$$

the root-mean-square (RMS) level of a signal reflects the signal energy.

C. Zero Crossing Rate

$$Z_t = \frac{1}{2}\sum_{n=1}^{N} |sign(x[n]) - sign(x[n-1])|, \tag{3}$$

a measure of the noisiness of the signal. $x[n]$ is the time domain signal for frame $t$.

D. Spectral Rolloff

The frequency such that,

$$\sum_{n=1}^{R_t} M_t[n] = 0.85 \sum_{n=1}^{N} M_t[n], \tag{4}$$

a measure of the skewness of the spectral shape. $M_t[n]$ is the magnitude of the Fourier transform at frame $t$ and frequency bin $n$.

E. Spectral Centroid

$$C_t = \frac{\sum_{n=1}^{N} M_t[n]*n}{\sum_{n=1}^{N} M_t[n]}, \tag{5}$$

the center of gravity of the magnitude spectrum of a short-time Fourier transform.

F. Spectral Flux

$$F_t = \sum_{n=1}^{N} (N_t[n] - N_{t-1}[n])^2, \tag{6}$$

a measure of the amount of local spectral change. $N_t[n]$ and $N_{t-1}[n]$ are the normalized magnitude of the Fourier transform at frames $t$ and $t-1$.

**Supplementary Methods 2 - TOPSIS**

1. Data gathering: We created a decision matrix where each row represented a different method and each column represented a metric. We then did this for each speech mode.

2. Data normalization: We normalized the decision matrix so that the metrics could be compared on the same scale. Vector normalization was implemented by dividing each element by the square root of the sum of squares of all elements in its column (Euclidean norm). We used this formula for normalization:

$$r_{ij} = \frac{x_{ij}}{\sqrt{\sum_{i=1}^{m} x_{ij}^2}}$$

Where $r_{ij}$ is the normalized value for method $i$ and criterion $j$, and $x_{ij}$ is the original value.

3. Assigning weights to metrics and calculation of weighted normalized matrix: We assumed equal importance for each metric (the sum of all weights equals 1). The process was then followed by multiplying each element of the normalized decision matrix by the weight of the corresponding metric:

$$v_{ij} = w_{ij} \cdot r_{ij}$$

4. Ideal and Anti-Ideal Solutions: For each metric, we identified the best and worst values from the weighted normalized matrix. The best values formed the "ideal best" and the worst values formed the "ideal worst" solution. Note that "best" may typically mean highest or lowest depending on whether the metric is a benefit or a cost criterion—in this study, all machine learning metrics are to be maximized.

5. Calculate the separation measures: For each alternative methods, calculate the Euclidean distance from the weighted normalized scores to both the ideal best and ideal worst:

$$D_i^+ = \sqrt{\sum_{j=1}^{n} (v_{ij} - A_j^+)^2}$$

$$D_i^- = \sqrt{\sum_{j=1}^{n} (v_{ij} - A_j^-)^2}$$

6. Calculate relative closeness: For each method, calculate the relative closeness to the ideal solution using the separation from the ideal best and ideal worst. This is done by dividing the separation from the ideal worst by the sum of the separation from the ideal best and ideal worst:

$$C_i^* = \frac{D_i^-}{D_i^+ + D_i^-}$$

7. Rank the alternatives: The method with the highest $C_i^*$ is preferred. A higher relative closeness indicates a better performance according to the specified metrics and weights.

### Supplementary Tables

**Table S1**. MCDA TOPSIS.

| dataset | mode | accuracy | sens. | spec. | precision | F1 | MCC | AUC | TOPSIS | rank | method |
|---|---|---|---|---|---|---|---|---|---|---|---|
| English [17] | read text | 0.67 | 0.36 | 0.90 | 0.43 | 0.37 | 0.28 | 0.72 | 0.41 | 4 | w2v1-FE |

| | | | | | | | | | | | |
|---|---|---|---|---|---|---|---|---|---|---|---|
| | | 0.75 | 0.48 | 0.93 | 0.63 | 0.53 | 0.44 | 0.75 | 0.85 | 2 | w2v1-FEA |
| | | 0.63 | 0.33 | 0.85 | 0.32 | 0.30 | 0.19 | 0.67 | 0.18 | 5 | w2v2-FE |
| | | 0.59 | 0.28 | 0.83 | 0.35 | 0.30 | 0.13 | 0.55 | 0.07 | 6 | w2v2-LH |
| | | 0.71 | 0.58 | 0.81 | 0.58 | 0.54 | 0.41 | 0.84 | 0.88 | 1 | w2v2-FT |
| | | 0.64 | 0.49 | 0.75 | 0.51 | 0.47 | 0.25 | 0.65 | 0.51 | 3 | baselines |
| | dialogue | 0.68 | 0.41 | 0.89 | 0.38 | 0.38 | 0.30 | 0.71 | 0.66 | 4 | w2v1-FE |
| | | 0.66 | 0.52 | 0.81 | 0.47 | 0.46 | 0.32 | 0.69 | 0.79 | 3 | w2v1-FEA |
| | | 0.60 | 0.36 | 0.80 | 0.29 | 0.29 | 0.16 | 0.65 | 0.43 | 5 | w2v2-FE |
| | | 0.53 | 0.18 | 0.77 | 0.19 | 0.17 | -0.06 | 0.45 | 0.02 | 6 | w2v2-LH |
| | | 0.65 | 0.59 | 0.73 | 0.48 | 0.50 | 0.31 | 0.71 | 0.81 | 2 | w2v2-FT |
| | | 0.69 | 0.57 | 0.80 | 0.60 | 0.54 | 0.39 | 0.73 | 0.95 | 1 | baselines |
| PC-GITA [18] | read text | 0.74 | 0.66 | 0.82 | 0.81 | 0.70 | 0.51 | 0.73 | 0.81 | 3 | w2v1-FE |
| | | 0.76 | 0.71 | 0.80 | 0.80 | 0.73 | 0.53 | 0.83 | 0.89 | 2 | w2v1-FEA |
| | | 0.70 | 0.72 | 0.68 | 0.73 | 0.70 | 0.42 | 0.76 | 0.68 | 4 | w2v2-FE |
| | | 0.55 | 0.46 | 0.64 | 0.53 | 0.47 | 0.10 | 0.59 | 0.10 | 6 | w2v2-LH |
| | | 0.78 | 0.78 | 0.77 | 0.81 | 0.77 | 0.57 | 0.82 | 0.96 | 1 | w2v2-FT |
| | | 0.57 | 0.62 | 0.53 | 0.57 | 0.58 | 0.15 | 0.59 | 0.20 | 5 | baselines |
| | monologue | 0.73 | 0.66 | 0.79 | 0.77 | 0.69 | 0.47 | 0.74 | 0.77 | 4 | w2v1-FE |
| | | 0.76 | 0.71 | 0.81 | 0.80 | 0.74 | 0.54 | 0.82 | 0.93 | 2 | w2v1-FEA |
| | | 0.74 | 0.77 | 0.71 | 0.75 | 0.75 | 0.51 | 0.80 | 0.87 | 3 | w2v2-FE |
| | | 0.62 | 0.48 | 0.75 | 0.61 | 0.51 | 0.25 | 0.64 | 0.27 | 5 | w2v2-LH |
| | | 0.76 | 0.76 | 0.76 | 0.78 | 0.76 | 0.54 | 0.82 | 0.94 | 1 | w2v2-FT |
| | | 0.57 | 0.58 | 0.56 | 0.60 | 0.57 | 0.15 | 0.61 | 0.13 | 6 | baselines |
| | vowels | 0.66 | 0.62 | 0.69 | 0.70 | 0.63 | 0.34 | 0.70 | 0.75 | 3 | w2v1-FE |
| | | 0.61 | 0.59 | 0.62 | 0.64 | 0.59 | 0.22 | 0.66 | 0.43 | 4 | w2v1-FEA |
| | | 0.69 | 0.73 | 0.65 | 0.69 | 0.70 | 0.39 | 0.76 | 0.95 | 1 | w2v2-FE |
| | | 0.61 | 0.57 | 0.64 | 0.62 | 0.58 | 0.23 | 0.61 | 0.41 | 5 | w2v2-LH |
| | | 0.68 | 0.71 | 0.66 | 0.69 | 0.69 | 0.38 | 0.74 | 0.93 | 2 | w2v2-FT |
| | | 0.55 | 0.44 | 0.66 | 0.58 | 0.46 | 0.11 | 0.55 | 0.05 | 6 | baselines |
| [19] | vowels | 0.63 | 0.63 | 0.62 | 0.65 | 0.61 | 0.27 | 0.62 | 0.40 | 3 | w2v1-FE |
| | | 0.61 | 0.63 | 0.59 | 0.61 | 0.60 | 0.23 | 0.65 | 0.30 | 5 | w2v1-FEA |
| | | 0.70 | 0.74 | 0.66 | 0.70 | 0.70 | 0.42 | 0.75 | 0.89 | 1 | w2v2-FE |
| | | 0.61 | 0.49 | 0.73 | 0.54 | 0.49 | 0.23 | 0.64 | 0.20 | 6 | w2v2-LH |
| | | 0.68 | 0.67 | 0.68 | 0.70 | 0.66 | 0.38 | 0.73 | 0.78 | 2 | w2v2-FT |
| | | 0.61 | 0.49 | 0.73 | 0.70 | 0.54 | 0.25 | 0.66 | 0.30 | 4 | baselines |

w2v2-FT: first transformer layer; w2v2-LH: last hidden layer; w2v2-FE: feature extraction; w2v1-FE: feature extraction; w2v1-FEA: feature extraction and aggregation

**Table S2.** Classifier performance metrics for read text from the English dataset, which included 21 HC and 16 PD samples in an imbalanced distribution. Results reported for the 30 PCA components.

| | classifier | accuracy | sens. | spec. | precision | F1 | MCC | AUC |
|---|---|---|---|---|---|---|---|---|
| **wav2vec 1.0 feature extractor** | DT | 0.54 | 0.35 | 0.65 | 0.25 | 0.28 | -0.02 | 0.50 |
| | KNN | 0.73 | 0.35 | 1.00 | 0.50 | 0.40 | 0.37 | 0.90 |
| | LR | 0.58 | 0.00 | 1.00 | 0.00 | 0.00 | 0.00 | 0.85 |
| | NB | 0.53 | 0.45 | 0.58 | 0.47 | 0.43 | 0.07 | 0.60 |
| | RF | 0.56 | 0.30 | 0.77 | 0.30 | 0.28 | 0.07 | 0.55 |
| | SVM | 0.58 | 0.00 | 1.00 | 0.00 | 0.00 | 0.00 | 0.33 |
| | **mean ± std** | **0.59 ± 0.07** | **0.24 ± 0.19** | **0.83 ± 0.19** | **0.25 ± 0.22** | **0.23 ± 0.19** | **0.08 ± 0.15** | **0.62 ± 0.22** |
| **wav2vec 1.0 feature extractor + aggregator** | DT | 0.40 | 0.15 | 0.57 | 0.20 | 0.17 | -0.30 | 0.36 |
| | KNN | 0.71 | 0.30 | 1.00 | 0.50 | 0.37 | 0.33 | 0.73 |
| | LR | 0.74 | 0.45 | 0.95 | 0.60 | 0.50 | 0.42 | 0.73 |
| | NB | 0.67 | 0.40 | 0.82 | 0.50 | 0.43 | 0.23 | 0.66 |
| | RF | 0.57 | 0.30 | 0.75 | 0.33 | 0.28 | 0.06 | 0.48 |
| | SVM | 0.73 | 0.35 | 1.00 | 0.50 | 0.40 | 0.37 | 0.75 |
| | **mean ± std** | **0.64 ± 0.13** | **0.33 ± 0.10** | **0.85 ± 0.17** | **0.44 ± 0.15** | **0.36 ± 0.12** | **0.19 ± 0.27** | **0.62 ± 0.16** |
| **wav2vec 2.0 feature extractor** | DT | 0.48 | 0.35 | 0.58 | 0.30 | 0.30 | -0.06 | 0.47 |
| | KNN | 0.66 | 0.20 | 1.00 | 0.30 | 0.23 | 0.22 | 0.67 |
| | LR | 0.58 | 0.00 | 1.00 | 0.00 | 0.00 | 0.00 | 0.75 |
| | NB | 0.68 | 0.65 | 0.72 | 0.55 | 0.58 | 0.37 | 0.82 |
| | RF | 0.58 | 0.35 | 0.77 | 0.30 | 0.30 | 0.11 | 0.72 |
| | SVM | 0.58 | 0.00 | 1.00 | 0.00 | 0.00 | 0.00 | 0.38 |
| | **mean ± std** | **0.59 ± 0.07** | **0.26 ± 0.25** | **0.84 ± 0.18** | **0.24 ± 0.21** | **0.24 ± 0.22** | **0.11 ± 0.16** | **0.63 ± 0.17** |
| **wav2vec 2.0 last hidden layer** | DT | 0.79 | 0.75 | 0.87 | 0.77 | 0.71 | 0.64 | 0.81 |
| | KNN | 0.66 | 0.55 | 0.78 | 0.60 | 0.53 | 0.36 | 0.66 |
| | LR | 0.58 | 0.00 | 1.00 | 0.00 | 0.00 | 0.00 | 0.38 |
| | NB | 0.79 | 0.75 | 0.80 | 0.65 | 0.68 | 0.55 | 0.85 |
| | RF | 0.79 | 0.55 | 0.95 | 0.67 | 0.58 | 0.53 | 0.68 |
| | SVM | 0.58 | 0.00 | 1.00 | 0.00 | 0.00 | 0.00 | 0.43 |
| | **mean ± std** | **0.70 ± 0.11** | **0.43 ± 0.35** | **0.90 ± 0.10** | **0.45 ± 0.35** | **0.42 ± 0.33** | **0.35 ± 0.28** | **0.64 ± 0.19** |
| **wav2vec 2.0 first transformer layer** | DT | 0.56 | 0.55 | 0.60 | 0.48 | 0.48 | 0.16 | 0.58 |
| | KNN | 0.61 | 0.15 | 0.95 | 0.20 | 0.17 | 0.10 | 0.76 |
| | LR | 0.73 | 0.65 | 0.80 | 0.62 | 0.60 | 0.47 | 0.95 |
| | NB | 0.76 | 0.60 | 0.85 | 0.62 | 0.60 | 0.47 | 0.80 |
| | RF | 0.53 | 0.30 | 0.75 | 0.28 | 0.27 | 0.06 | 0.63 |
| | SVM | 0.76 | 0.65 | 0.85 | 0.67 | 0.63 | 0.52 | 0.90 |

| | mean ± std | **0.66 ± 0.10** | **0.48 ± 0.21** | **0.80 ± 0.12** | **0.48 ± 0.19** | **0.46 ± 0.20** | **0.30 ± 0.21** | **0.77 ± 0.15** |
|---|---|---|---|---|---|---|---|---|

**Table S3.** Classifier performance metrics for dialogue from the English dataset, which included 21 HC and 15 PD samples in an imbalanced distribution. Results reported for the 30 PCA components.

| | classifier | accuracy | sens. | spec. | precision | F1 | MCC | AUC |
|---|---|---|---|---|---|---|---|---|
| **wav2vec 1.0 feature extractor** | DT | 0.51 | 0.40 | 0.58 | 0.25 | 0.30 | -0.01 | 0.49 |
| | KNN | 0.72 | 0.45 | 0.92 | 0.50 | 0.47 | 0.37 | 0.61 |
| | LR | 0.59 | 0.00 | 1.00 | 0.00 | 0.00 | 0.00 | 0.82 |
| | NB | 0.63 | 0.50 | 0.70 | 0.42 | 0.43 | 0.22 | 0.58 |
| | RF | 0.51 | 0.10 | 0.77 | 0.10 | 0.10 | -0.14 | 0.43 |
| | SVM | 0.59 | 0.00 | 1.00 | 0.00 | 0.00 | 0.00 | 0.62 |
| | **mean ± std** | **0.59 ± 0.08** | **0.24 ± 0.23** | **0.83 ± 0.17** | **0.21 ± 0.21** | **0.22 ± 0.21** | **0.07 ± 0.18** | **0.59 ± 0.13** |
| **wav2vec 1.0 feature extractor + aggregator** | DT | 0.50 | 0.45 | 0.57 | 0.28 | 0.33 | 0.00 | 0.51 |
| | KNN | 0.68 | 0.55 | 0.82 | 0.50 | 0.50 | 0.35 | 0.62 |
| | LR | 0.72 | 0.50 | 0.92 | 0.50 | 0.48 | 0.42 | 0.79 |
| | NB | 0.62 | 0.35 | 0.75 | 0.43 | 0.36 | 0.14 | 0.68 |
| | RF | 0.53 | 0.10 | 0.80 | 0.20 | 0.13 | -0.08 | 0.55 |
| | SVM | 0.72 | 0.50 | 0.92 | 0.50 | 0.48 | 0.42 | 0.69 |
| | **mean ± std** | **0.63 ± 0.10** | **0.41 ± 0.17** | **0.79 ± 0.13** | **0.40 ± 0.13** | **0.38 ± 0.14** | **0.21 ± 0.22** | **0.64 ± 0.10** |
| **wav2vec 2.0 feature extractor** | DT | 0.78 | 0.80 | 0.78 | 0.65 | 0.70 | 0.58 | 0.79 |
| | KNN | 0.66 | 0.40 | 0.87 | 0.45 | 0.40 | 0.27 | 0.68 |
| | LR | 0.59 | 0.00 | 1.00 | 0.00 | 0.00 | 0.00 | 0.72 |
| | NB | 0.50 | 0.25 | 0.67 | 0.22 | 0.21 | -0.10 | 0.42 |
| | RF | 0.59 | 0.30 | 0.82 | 0.28 | 0.27 | 0.11 | 0.52 |
| | SVM | 0.59 | 0.00 | 1.00 | 0.00 | 0.00 | 0.00 | 0.42 |
| | **mean ± std** | **0.62 ± 0.09** | **0.29 ± 0.30** | **0.86 ± 0.13** | **0.27 ± 0.26** | **0.26 ± 0.26** | **0.14 ± 0.25** | **0.59 ± 0.16** |
| **wav2vec 2.0 last hidden layer** | DT | 0.54 | 0.40 | 0.57 | 0.33 | 0.36 | -0.04 | 0.48 |
| | KNN | 0.44 | 0.10 | 0.67 | 0.13 | 0.11 | -0.24 | 0.38 |
| | LR | 0.59 | 0.00 | 1.00 | 0.00 | 0.00 | 0.00 | 0.30 |
| | NB | 0.63 | 0.35 | 0.80 | 0.42 | 0.36 | 0.17 | 0.59 |
| | RF | 0.68 | 0.40 | 0.85 | 0.50 | 0.43 | 0.26 | 0.58 |
| | SVM | 0.59 | 0.00 | 1.00 | 0.00 | 0.00 | 0.00 | 0.53 |
| | **mean ± std** | **0.58 ± 0.08** | **0.21 ± 0.20** | **0.81 ± 0.18** | **0.23 ± 0.22** | **0.21 ± 0.20** | **0.02 ± 0.17** | **0.48 ± 0.11** |
| **wav2vec 2.0 first transformer layer** | DT | 0.53 | 0.60 | 0.52 | 0.43 | 0.47 | 0.12 | 0.56 |
| | KNN | 0.68 | 0.55 | 0.82 | 0.53 | 0.51 | 0.37 | 0.66 |
| | LR | 0.67 | 0.55 | 0.77 | 0.50 | 0.52 | 0.30 | 0.73 |
| | NB | 0.36 | 0.15 | 0.47 | 0.12 | 0.13 | -0.41 | 0.37 |

| | | | | | | | | |
|---|---|---|---|---|---|---|---|---|
| | RF | 0.53 | 0.25 | 0.77 | 0.25 | 0.23 | 0.01 | 0.53 |
| | SVM | 0.60 | 0.55 | 0.67 | 0.55 | 0.52 | 0.22 | 0.73 |
| | **mean ± std** | **0.56 ± 0.12** | **0.44 ± 0.19** | **0.67 ± 0.14** | **0.40 ± 0.18** | **0.40 ± 0.17** | **0.10 ± 0.28** | **0.59 ± 0.14** |

**Table S4.** Classifier performance metrics for read text from the PC-GITA dataset, which included 50 HC and 50 PD samples in a balanced distribution. Results reported for the 30 PCA components.

| | **classifier** | **accuracy** | **sens.** | **spec.** | **precision** | **F1** | **MCC** | **AUC** |
|---|---|---|---|---|---|---|---|---|
| **wav2vec 1.0 feature extractor** | DT | 0.65 | 0.74 | 0.56 | 0.64 | 0.67 | 0.30 | 0.65 |
| | KNN | 0.77 | 0.62 | 0.92 | 0.91 | 0.71 | 0.58 | 0.82 |
| | LR | 0.69 | 0.58 | 0.80 | 0.76 | 0.64 | 0.40 | 0.75 |
| | NB | 0.67 | 0.66 | 0.68 | 0.71 | 0.67 | 0.36 | 0.77 |
| | RF | 0.74 | 0.74 | 0.74 | 0.78 | 0.75 | 0.50 | 0.80 |
| | SVM | 0.66 | 0.50 | 0.82 | 0.75 | 0.59 | 0.35 | 0.34 |
| | **mean ± std** | **0.70 ± 0.05** | **0.64 ± 0.09** | **0.75 ± 0.12** | **0.76 ± 0.09** | **0.67 ± 0.05** | **0.41 ± 0.11** | **0.69 ± 0.18** |
| **wav2vec 1.0 feature extractor + aggregator** | DT | 0.63 | 0.70 | 0.56 | 0.62 | 0.65 | 0.27 | 0.63 |
| | KNN | 0.77 | 0.62 | 0.92 | 0.87 | 0.71 | 0.57 | 0.83 |
| | LR | 0.75 | 0.68 | 0.82 | 0.80 | 0.71 | 0.52 | 0.87 |
| | NB | 0.68 | 0.68 | 0.68 | 0.68 | 0.67 | 0.37 | 0.74 |
| | RF | 0.76 | 0.80 | 0.72 | 0.74 | 0.76 | 0.54 | 0.80 |
| | SVM | 0.81 | 0.76 | 0.86 | 0.86 | 0.78 | 0.65 | 0.89 |
| | **mean ± std** | **0.73 ± 0.07** | **0.71 ± 0.06** | **0.76 ± 0.13** | **0.76 ± 0.10** | **0.71 ± 0.05** | **0.49 ± 0.14** | **0.79 ± 0.10** |
| **wav2vec 2.0 feature extractor** | DT | 0.64 | 0.58 | 0.70 | 0.65 | 0.60 | 0.28 | 0.64 |
| | KNN | 0.75 | 0.66 | 0.84 | 0.83 | 0.71 | 0.53 | 0.82 |
| | LR | 0.68 | 0.76 | 0.60 | 0.70 | 0.71 | 0.37 | 0.77 |
| | NB | 0.70 | 0.68 | 0.72 | 0.75 | 0.70 | 0.42 | 0.76 |
| | RF | 0.72 | 0.68 | 0.76 | 0.78 | 0.70 | 0.46 | 0.80 |
| | SVM | 0.69 | 0.78 | 0.60 | 0.71 | 0.73 | 0.39 | 0.76 |
| | **mean ± std** | **0.70 ± 0.04** | **0.69 ± 0.07** | **0.70 ± 0.09** | **0.74 ± 0.06** | **0.69 ± 0.04** | **0.41 ± 0.08** | **0.76 ± 0.06** |
| **wav2vec 2.0 last hidden layer** | DT | 0.74 | 0.72 | 0.76 | 0.78 | 0.72 | 0.51 | 0.74 |
| | KNN | 0.56 | 0.48 | 0.64 | 0.58 | 0.51 | 0.13 | 0.63 |
| | LR | 0.41 | 0.34 | 0.48 | 0.41 | 0.36 | -0.19 | 0.38 |
| | NB | 0.71 | 0.66 | 0.76 | 0.75 | 0.69 | 0.43 | 0.80 |
| | RF | 0.77 | 0.80 | 0.74 | 0.78 | 0.76 | 0.58 | 0.88 |
| | SVM | 0.45 | 0.18 | 0.72 | 0.33 | 0.21 | -0.12 | 0.56 |
| | **mean ± std** | **0.61 ± 0.16** | **0.53 ± 0.24** | **0.68 ± 0.11** | **0.61 ± 0.20** | **0.54 ± 0.22** | **0.22 ± 0.33** | **0.66 ± 0.18** |
| **wav2vec 2.0 first transformer** | DT | 0.65 | 0.64 | 0.66 | 0.67 | 0.64 | 0.32 | 0.65 |
| | KNN | 0.76 | 0.68 | 0.84 | 0.84 | 0.73 | 0.55 | 0.78 |

| | | | | | | | | |
|---|---|---|---|---|---|---|---|---|
| **layer** | LR | 0.81 | 0.78 | 0.84 | 0.86 | 0.81 | 0.64 | 0.83 |
| | NB | 0.74 | 0.74 | 0.74 | 0.77 | 0.74 | 0.50 | 0.84 |
| | RF | 0.75 | 0.78 | 0.72 | 0.77 | 0.76 | 0.51 | 0.77 |
| | SVM | 0.79 | 0.80 | 0.78 | 0.82 | 0.79 | 0.60 | 0.80 |
| | **mean ± std** | **0.75 ± 0.06** | **0.74 ± 0.06** | **0.76 ± 0.07** | **0.79 ± 0.07** | **0.74 ± 0.06** | **0.52 ± 0.11** | **0.78 ± 0.07** |

**Table S5.** Classifier performance metrics for monologue from the PC-GITA dataset, which included 50 HC and 50 PD samples in a balanced distribution. Results reported for the 30 PCA components.

| | **classifier** | **accuracy** | **sens.** | **spec.** | **precision** | **F1** | **MCC** | **AUC** |
|---|---|---|---|---|---|---|---|---|
| **wav2vec 1.0 feature extractor** | DT | 0.65 | 0.64 | 0.66 | 0.66 | 0.64 | 0.32 | 0.65 |
| | KNN | 0.75 | 0.68 | 0.82 | 0.74 | 0.70 | 0.50 | 0.83 |
| | LR | 0.69 | 0.62 | 0.76 | 0.73 | 0.65 | 0.39 | 0.80 |
| | NB | 0.81 | 0.84 | 0.78 | 0.80 | 0.82 | 0.62 | 0.86 |
| | RF | 0.77 | 0.72 | 0.82 | 0.83 | 0.74 | 0.57 | 0.86 |
| | SVM | 0.65 | 0.52 | 0.78 | 0.73 | 0.58 | 0.33 | 0.39 |
| | **mean ± std** | **0.72 ± 0.07** | **0.67 ± 0.11** | **0.77 ± 0.06** | **0.75 ± 0.06** | **0.69 ± 0.08** | **0.45 ± 0.13** | **0.73 ± 0.19** |
| **wav2vec 1.0 feature extractor + aggregator** | DT | 0.67 | 0.68 | 0.66 | 0.68 | 0.66 | 0.35 | 0.67 |
| | KNN | 0.76 | 0.72 | 0.80 | 0.80 | 0.75 | 0.54 | 0.80 |
| | LR | 0.77 | 0.74 | 0.80 | 0.81 | 0.76 | 0.56 | 0.87 |
| | NB | 0.73 | 0.70 | 0.76 | 0.75 | 0.71 | 0.48 | 0.80 |
| | RF | 0.69 | 0.68 | 0.70 | 0.70 | 0.67 | 0.41 | 0.79 |
| | SVM | 0.79 | 0.74 | 0.84 | 0.84 | 0.77 | 0.61 | 0.90 |
| | **mean ± std** | **0.74 ± 0.05** | **0.71 ± 0.03** | **0.76 ± 0.07** | **0.76 ± 0.06** | **0.72 ± 0.05** | **0.49 ± 0.10** | **0.81 ± 0.08** |
| **wav2vec 2.0 feature extractor** | DT | 0.66 | 0.62 | 0.70 | 0.67 | 0.64 | 0.33 | 0.66 |
| | KNN | 0.78 | 0.74 | 0.82 | 0.83 | 0.76 | 0.58 | 0.83 |
| | LR | 0.69 | 0.74 | 0.64 | 0.68 | 0.70 | 0.40 | 0.78 |
| | NB | 0.73 | 0.70 | 0.76 | 0.76 | 0.72 | 0.47 | 0.80 |
| | RF | 0.74 | 0.76 | 0.72 | 0.75 | 0.75 | 0.49 | 0.81 |
| | SVM | 0.69 | 0.80 | 0.58 | 0.67 | 0.72 | 0.41 | 0.79 |
| | **mean ± std** | **0.72 ± 0.04** | **0.73 ± 0.06** | **0.70 ± 0.09** | **0.73 ± 0.06** | **0.71 ± 0.04** | **0.45 ± 0.09** | **0.78 ± 0.06** |
| **wav2vec 2.0 last hidden layer** | DT | 0.61 | 0.64 | 0.58 | 0.64 | 0.63 | 0.23 | 0.61 |
| | KNN | 0.61 | 0.54 | 0.68 | 0.65 | 0.56 | 0.24 | 0.66 |
| | LR | 0.56 | 0.52 | 0.60 | 0.59 | 0.52 | 0.14 | 0.63 |
| | NB | 0.76 | 0.78 | 0.74 | 0.76 | 0.75 | 0.55 | 0.89 |
| | RF | 0.74 | 0.68 | 0.80 | 0.80 | 0.72 | 0.50 | 0.80 |
| | SVM | 0.53 | 0.08 | 0.98 | 0.35 | 0.13 | 0.10 | 0.39 |
| | **mean ± std** | **0.64 ± 0.09** | **0.54 ± 0.24** | **0.73 ± 0.15** | **0.63 ± 0.16** | **0.55 ± 0.23** | **0.29 ± 0.19** | **0.66 ± 0.17** |

| | | | | | | | | |
|---|---|---|---|---|---|---|---|---|
| **wav2vec 2.0 first transformer layer** | DT | 0.54 | 0.56 | 0.52 | 0.54 | 0.54 | 0.07 | 0.54 |
| | KNN | 0.75 | 0.72 | 0.78 | 0.78 | 0.74 | 0.52 | 0.83 |
| | LR | 0.80 | 0.80 | 0.80 | 0.82 | 0.80 | 0.62 | 0.90 |
| | NB | 0.75 | 0.82 | 0.68 | 0.72 | 0.77 | 0.51 | 0.82 |
| | RF | 0.74 | 0.76 | 0.72 | 0.74 | 0.75 | 0.48 | 0.79 |
| | SVM | 0.77 | 0.78 | 0.76 | 0.81 | 0.77 | 0.57 | 0.88 |
| | **mean ± std** | **0.73 ± 0.09** | **0.74 ± 0.09** | **0.71 ± 0.10** | **0.74 ± 0.10** | **0.73 ± 0.09** | **0.46 ± 0.20** | **0.79 ± 0.13** |

**Table S6.** Classifier performance metrics for vowel dataset from the PC-GITA dataset, which included 50 HC and 50 PD samples in a balanced distribution. Results reported for the 30 PCA components.

| | **classifier** | **accuracy** | **sens.** | **spec.** | **precision** | **F1** | **MCC** | **AUC** |
|---|---|---|---|---|---|---|---|---|
| **wav2vec 1.0 feature extractor** | DT | 0.59 | 0.64 | 0.54 | 0.60 | 0.61 | 0.18 | 0.59 |
| | KNN | 0.62 | 0.64 | 0.60 | 0.63 | 0.62 | 0.25 | 0.65 |
| | LR | 0.65 | 0.64 | 0.66 | 0.67 | 0.64 | 0.32 | 0.71 |
| | NB | 0.63 | 0.68 | 0.58 | 0.63 | 0.65 | 0.26 | 0.66 |
| | RF | 0.64 | 0.72 | 0.56 | 0.63 | 0.66 | 0.29 | 0.69 |
| | SVM | 0.61 | 0.44 | 0.78 | 0.72 | 0.51 | 0.26 | 0.73 |
| | **mean ± std** | **0.62 ± 0.02** | **0.63 ± 0.10** | **0.62 ± 0.09** | **0.65 ± 0.05** | **0.61 ± 0.05** | **0.26 ± 0.05** | **0.67 ± 0.05** |
| **wav2vec 1.0 feature extractor + aggregator** | DT | 0.44 | 0.42 | 0.46 | 0.40 | 0.40 | -0.13 | 0.44 |
| | KNN | 0.53 | 0.60 | 0.46 | 0.53 | 0.55 | 0.04 | 0.57 |
| | LR | 0.58 | 0.54 | 0.62 | 0.62 | 0.55 | 0.17 | 0.68 |
| | NB | 0.63 | 0.54 | 0.72 | 0.65 | 0.57 | 0.27 | 0.67 |
| | RF | 0.56 | 0.52 | 0.60 | 0.58 | 0.53 | 0.13 | 0.59 |
| | SVM | 0.55 | 0.50 | 0.60 | 0.58 | 0.52 | 0.11 | 0.69 |
| | **mean ± std** | **0.55 ± 0.06** | **0.52 ± 0.06** | **0.58 ± 0.10** | **0.56 ± 0.09** | **0.52 ± 0.06** | **0.10 ± 0.13** | **0.61 ± 0.10** |
| **wav2vec 2.0 feature extractor** | DT | 0.53 | 0.60 | 0.46 | 0.53 | 0.55 | 0.07 | 0.53 |
| | KNN | 0.74 | 0.76 | 0.72 | 0.76 | 0.75 | 0.49 | 0.80 |
| | LR | 0.71 | 0.76 | 0.66 | 0.71 | 0.73 | 0.43 | 0.83 |
| | NB | 0.76 | 0.76 | 0.76 | 0.78 | 0.76 | 0.53 | 0.80 |
| | RF | 0.69 | 0.76 | 0.62 | 0.69 | 0.71 | 0.39 | 0.81 |
| | SVM | 0.73 | 0.72 | 0.74 | 0.77 | 0.73 | 0.47 | 0.80 |
| | **mean ± std** | **0.69 ± 0.08** | **0.73 ± 0.06** | **0.66 ± 0.11** | **0.71 ± 0.09** | **0.71 ± 0.08** | **0.40 ± 0.17** | **0.76 ± 0.11** |
| **wav2vec 2.0 last hidden layer** | DT | 0.63 | 0.58 | 0.68 | 0.62 | 0.59 | 0.26 | 0.63 |
| | KNN | 0.68 | 0.72 | 0.64 | 0.71 | 0.69 | 0.38 | 0.67 |
| | LR | 0.59 | 0.48 | 0.70 | 0.61 | 0.53 | 0.19 | 0.64 |
| | NB | 0.63 | 0.62 | 0.64 | 0.62 | 0.61 | 0.27 | 0.67 |
| | RF | 0.69 | 0.70 | 0.68 | 0.70 | 0.69 | 0.39 | 0.78 |

| | | | | | | | | |
|---|---|---|---|---|---|---|---|---|
| | SVM | 0.56 | 0.36 | 0.76 | 0.62 | 0.43 | 0.14 | 0.42 |
| | **mean ± std** | **0.63 ± 0.05** | **0.58 ± 0.14** | **0.68 ± 0.04** | **0.65 ± 0.04** | **0.59 ± 0.10** | **0.27 ± 0.10** | **0.63 ± 0.12** |
| **wav2vec 2.0 first transformer layer** | DT | 0.62 | 0.74 | 0.50 | 0.60 | 0.65 | 0.28 | 0.62 |
| | KNN | 0.73 | 0.76 | 0.70 | 0.72 | 0.74 | 0.47 | 0.79 |
| | LR | 0.70 | 0.72 | 0.68 | 0.70 | 0.70 | 0.41 | 0.78 |
| | NB | 0.72 | 0.74 | 0.70 | 0.72 | 0.72 | 0.45 | 0.72 |
| | RF | 0.68 | 0.70 | 0.66 | 0.68 | 0.68 | 0.36 | 0.76 |
| | SVM | 0.74 | 0.78 | 0.70 | 0.74 | 0.75 | 0.50 | 0.78 |
| | **mean ± std** | **0.70 ± 0.04** | **0.74 ± 0.03** | **0.66 ± 0.08** | **0.69 ± 0.05** | **0.71 ± 0.04** | **0.41 ± 0.08** | **0.74 ± 0.07** |

**Table S7.** Classifier performance metrics for vowel dataset from the U.S.-based dataset, which included 41 HC and 40 PD samples in a balanced distribution. Results reported for the 30 PCA components.

| | **classifier** | **accuracy** | **sens.** | **spec.** | **precision** | **F1** | **MCC** | **AUC** |
|---|---|---|---|---|---|---|---|---|
| **wav2vec 1.0 feature extractor** | DT | 0.52 | 0.60 | 0.44 | 0.52 | 0.54 | 0.06 | 0.52 |
| | KNN | 0.54 | 0.70 | 0.40 | 0.58 | 0.59 | 0.12 | 0.61 |
| | LR | 0.63 | 0.65 | 0.62 | 0.64 | 0.63 | 0.28 | 0.68 |
| | NB | 0.67 | 0.63 | 0.71 | 0.71 | 0.64 | 0.35 | 0.68 |
| | RF | 0.65 | 0.65 | 0.65 | 0.68 | 0.65 | 0.32 | 0.69 |
| | SVM | 0.63 | 0.63 | 0.64 | 0.66 | 0.61 | 0.29 | 0.37 |
| | **mean ± std** | **0.61 ± 0.06** | **0.64 ± 0.03** | **0.57 ± 0.13** | **0.63 ± 0.07** | **0.61 ± 0.04** | **0.24 ± 0.12** | **0.59 ± 0.13** |
| **wav2vec 1.0 feature extractor + aggregator** | DT | 0.50 | 0.55 | 0.43 | 0.48 | 0.50 | -0.04 | 0.49 |
| | KNN | 0.69 | 0.70 | 0.68 | 0.74 | 0.69 | 0.41 | 0.72 |
| | LR | 0.59 | 0.65 | 0.54 | 0.63 | 0.61 | 0.20 | 0.65 |
| | NB | 0.53 | 0.60 | 0.46 | 0.53 | 0.56 | 0.05 | 0.50 |
| | RF | 0.61 | 0.63 | 0.59 | 0.55 | 0.58 | 0.20 | 0.65 |
| | SVM | 0.64 | 0.68 | 0.61 | 0.65 | 0.65 | 0.29 | 0.71 |
| | **mean ± std** | **0.59 ± 0.07** | **0.63 ± 0.05** | **0.55 ± 0.09** | **0.60 ± 0.09** | **0.60 ± 0.07** | **0.19 ± 0.16** | **0.62 ± 0.10** |
| **wav2vec 2.0 feature extractor** | DT | 0.64 | 0.65 | 0.63 | 0.70 | 0.64 | 0.31 | 0.64 |
| | KNN | 0.65 | 0.73 | 0.58 | 0.68 | 0.66 | 0.34 | 0.68 |
| | LR | 0.71 | 0.75 | 0.68 | 0.72 | 0.72 | 0.45 | 0.80 |
| | NB | 0.62 | 0.55 | 0.68 | 0.60 | 0.54 | 0.25 | 0.68 |
| | RF | 0.67 | 0.68 | 0.66 | 0.68 | 0.65 | 0.36 | 0.74 |
| | SVM | 0.71 | 0.75 | 0.68 | 0.72 | 0.72 | 0.45 | 0.83 |
| | **mean ± std** | **0.67 ± 0.04** | **0.68 ± 0.08** | **0.65 ± 0.04** | **0.68 ± 0.04** | **0.65 ± 0.07** | **0.36 ± 0.08** | **0.73 ± 0.07** |
| **wav2vec 2.0 last hidden layer** | DT | 0.63 | 0.68 | 0.58 | 0.67 | 0.65 | 0.28 | 0.63 |
| | KNN | 0.64 | 0.75 | 0.53 | 0.62 | 0.67 | 0.29 | 0.72 |
| | LR | 0.62 | 0.40 | 0.83 | 0.59 | 0.46 | 0.24 | 0.72 |

| | | | | | | | | |
|---|---|---|---|---|---|---|---|---|
| | NB | 0.62 | 0.58 | 0.66 | 0.64 | 0.59 | 0.25 | 0.68 |
| | RF | 0.59 | 0.70 | 0.48 | 0.59 | 0.63 | 0.17 | 0.69 |
| | SVM | 0.51 | 0.00 | 1.00 | 0.00 | 0.00 | 0.00 | 0.29 |
| | **mean ± std** | **0.60 ± 0.05** | **0.52 ± 0.28** | **0.68 ± 0.20** | **0.52 ± 0.26** | **0.50 ± 0.26** | **0.20 ± 0.11** | **0.62 ± 0.17** |
| **wav2vec 2.0 first transformer layer** | DT | 0.62 | 0.63 | 0.61 | 0.64 | 0.62 | 0.25 | 0.62 |
| | KNN | 0.67 | 0.68 | 0.66 | 0.71 | 0.65 | 0.37 | 0.71 |
| | LR | 0.64 | 0.63 | 0.66 | 0.65 | 0.62 | 0.30 | 0.69 |
| | NB | 0.60 | 0.55 | 0.67 | 0.61 | 0.55 | 0.23 | 0.65 |
| | RF | 0.65 | 0.65 | 0.66 | 0.65 | 0.63 | 0.32 | 0.69 |
| | SVM | 0.60 | 0.55 | 0.66 | 0.63 | 0.55 | 0.23 | 0.69 |
| | **mean ± std** | **0.63 ± 0.03** | **0.61 ± 0.05** | **0.65 ± 0.02** | **0.65 ± 0.04** | **0.60 ± 0.04** | **0.28 ± 0.06** | **0.68 ± 0.03** |
